# Daily Paced Breathing Sessions Induce Left Orbitofrontal Volume Changes Linked to Cognitive Outcomes

**DOI:** 10.64898/2026.03.02.26347452

**Authors:** Hyun Joo Yoo, Andy Jeesu Kim, Martin J. Dahl, Kalekirstos Alemu, Kaoru Nashiro, Christine Cho, Noah Mercer, Paul Choi, Hye Rynn, J. Lee, Jungwon Min, Nicole F. Rose, Julian F. Thayer, Mara Mather

**Affiliations:** Leonard Davis School of Gerontology, University of Southern California; Center for Lifespan Psychology, Max Planck Institute for Human Development; Max Planck UCL Centre for Computational Psychiatry and Ageing Research; Lumos Labs, Inc.; University of California, Irvine; Department of Psychology, University of Southern California; Department of Biomedical Engineering, University of Southern California

**Keywords:** breathing, heart rate variability, orbitofrontal cortex, structural plasticity, cognitive control, aging

## Abstract

Oscillatory coupling between respiration, heart rate, and cortical function is fundamental to physiological regulation yet remains poorly characterized in humans. Diminished respiratory heart rate variability (RespHRV)—the rhythmic heart rate modulation accompanying respiration—has emerged as a transdiagnostic biomarker of mental and physical health, reduced in anxiety, depression, cardiovascular disease, and aging (Beauchaine & Thayer, 2015; Menuet & Gourine et al., 2025). However, the cortical substrates that coordinate rhythmic cardiovascular–respiratory coupling are not well understood.

Our current findings highlight the involvement of the left orbitofrontal cortex (OFC) in oscillatory cardiorespiratory dynamics. In adults aged 50–70 (N = 55; mean age = 60.1 ± 6.0 years; 29 female), across both a slow-paced breathing condition and a random-paced breathing condition, greater heart rate oscillatory power during 9-week breathing training sessions predicted OFC volume increases. OFC changes were most strongly linked with upper low-frequency range power during practice (0.09–0.13 Hz; *p* < 0.005, cluster-corrected) but were not tightly constrained by precise breathing frequency. These effects covaried with improved attentional and executive performance, including reduced pupil responses to distractors and enhanced working-memory and associative-memory scores.

Our findings identify the orbitofrontal cortex as a key site of cortical plasticity linked to rhythmic cardiovascular–respiratory engagement. By delineating how oscillatory body–brain coupling supports cognitive control–related processes, including attentional filtering and memory updating, this work bridges mechanistic neuroscience and translational intervention science, suggesting a frequency-general pathway through which simple breathing practices may enhance neurovisceral integration and cognitive resilience in aging.

**Summary:**

- Greater oscillatory heart rate power during breathing training, particularly within the upper low-frequency range (0.09–0.13 Hz), predicted increases in left orbitofrontal cortex (OFC) volume.
- OFC volume increases were associated with improved attentional and executive performance, including reduced pupil reactivity to distractors and enhanced working-memory and associative-memory scores.
- These findings suggest that rhythmic cardiovascular–respiratory coupling supports cortical plasticity and cognitive resilience, providing a frequency-general mechanism through which breathing practices enhance neurovisceral integration in aging.

## 1. Introduction

Cardiorespiratory rhythms are among the most fundamental oscillations in the human body. Variability in heart rate across the respiratory cycle—termed respiratory heart rate variability (RespHRV)—reflects the degree of synchronization between respiratory and cardiovascular control systems (Menuet et al., 2025). Oscillations in heart rate at the breathing frequency are a dominant contributor to typical measures of heart rate variability (HRV), particularly those oscillations in the high-frequency range (Ritz, 2024). High-frequency HRV (HF-HRV), typically defined as power in the 0.15–0.40 Hz range, has been identified as a transdiagnostic biomarker of vulnerability across disorders, including depression, anxiety, cardiovascular disease, and neurodegenerative conditions (Beauchaine & Thayer, 2015; Chalmers et al., 2014). Evidence from occupational and population-level studies likewise links lower vagally mediated HRV to chronic stress and increased health risk, underscoring its role as a systems-level marker of resilience(Jarczok et al., 2013, 2022) Conversely, higher high-frequency HRV is linked to improved emotion regulation, stress recovery, and cognitive flexibility (Williams et al., 2019). Understanding the neural correlates of rhythmic cardio–respiratory coupling has thus become a central challenge for neuroscience and behavioral medicine alike.

Emerging evidence suggests that prefrontal regions—especially the orbitofrontal cortex (OFC)—play a central role in integrating autonomic and cognitive processes. Structural and functional neuroimaging studies have associated greater HRV during rest with greater left orbitofrontal cortical thickness (Koenig et al., 2021; Yoo et al., 2018), and increasing RespHRV using paced breathing and biofeedback has been found to increase left orbitofrontal structural volume compared with biofeedback to reduce HRV (Yoo et al., 2022). More generally, other neuroimaging studies have linked higher HRV to both greater structural integrity and functional activation in prefrontal regions involved in emotion regulation and autonomic control, including the medial prefrontal cortex (PFC), anterior cingulate cortex, and lateral OFC (Åhs et al., 2009; Chang et al., 2013; Critchley et al., 2003; Lane et al., 2009; Matthews et al., 2004; Smith et al., 2015; Thayer et al., 2009; Thayer & Lane, 2000; Yoo et al., 2018). Systematic reviews of the neuroimaging literature further indicate that heart rate variability measures are robustly associated with both structural and functional properties of multiple brain regions implicated in autonomic regulation, including prefrontal and limbic cortices (Matusik et al., 2023). Theoretical models propose that prefrontal–autonomic networks maintain homeostatic balance by coordinating attention, emotion, and visceral regulation (Critchley et al., 2003; Lane et al., 2009). Yet, the processes linking HRV and cortical structure remain largely unknown. The previous randomized controlled trial we conducted (“HRV-ER”; Yoo et al., 2022) suggests several possibilities. In that study, we assigned participants to either an Osc+ or Osc- condition. In the Osc+ condition, they completed five weeks of daily sessions involving slow paced breathing (around 10s/breath or 0.1 Hz) while getting positive biofeedback when heart rate oscillations were high. In the Osc- condition, they got no breathing instructions while getting positive biofeedback when heart rate oscillations were low. The two conditions had opposing effects on left orbitofrontal cortical volume, with the Osc+ condition increasing and the Osc-condition decreasing it, in both younger and older adults. These findings indicate that something about the HRV biofeedback practice influenced orbitofrontal structural volume, but it is not clear whether it was the: 1) conscious attempts to implement the biofeedback goals; 2) the specific frequency of respiratory-heart rate oscillations; or 3) the extent of heart rate oscillation during practice sessions that was the critical factor in producing the structural changes. Importantly, emerging evidence suggests that distinct frequency components of HRV may differentially relate to cortical control networks, underscoring the importance of frequency-resolved analyses when examining brain–autonomic relationships (Schneider & Schwerdtfeger, 2020).

In the current study, we used data from our Heart rate and Breathing Effects on Attention and Memory (‘HeartBEAM’) clinical trial (Nashiro et al., 2024) to investigate whether paced breathing induces structural plasticity in the orbitofrontal cortex (OFC) and whether such plasticity relates to changes in cognitive performance. Primary outcomes in this trial have been reported elsewhere (Nashiro et al., 2026). Adults aged 50–70 completed nine weeks of paced breathing training, randomized to either a condition with individualized calibration of breathing frequencies to maximize low-frequency heart rate variability (LF-HRV; typically defined as power in the 0.04–0.15 Hz range), or to a condition where breathing frequency was paced to correspond with typical breathing rates (4-6 s/breath) and inhalation and exhalation lengths differed randomly within a small range. The frequency of breathing may matter because it is hypothesized that HRV biofeedback enhances heart rate oscillations by synchronizing two physiological rhythms: respiration and baroreflex activity (Lehrer, 2013; Vaschillo et al., 2006). The baroreflex, a closed-loop system that regulates blood pressure via heart rate adjustments, exhibits a delay of approximately 4–6.5 seconds between changes in blood pressure and the resulting heart rate response (Ottesen, 1997; Vaschillo et al., 2011). This delay gives rise to heart rate oscillations within a frequency band of roughly 0.075–0.125 Hz (Mather & Thayer, 2018; Vaschillo et al., 2006). When breathing is paced within this same range, typically around six breaths per minute, these oscillations have been hypothesized to be amplified through resonance (Lehrer, 2013; Vaschillo et al., 2006), a non-linear interaction that enhances autonomic regulation. The low-frequency band of HRV is believed to reflect baroreflex-mediated oscillations involving both sympathetic and parasympathetic influences (Goldstein et al., 2011; Reyes del Paso et al., 2013), however when people breathe at a frequency within this range, the increased oscillations in heart rate at the breathing frequency are almost entirely driven by parasympathetic activity and not by sympathetic activity (Kromenacker et al., 2018). Most HRV biofeedback protocols calibrate breathing rates to an individual’s resonance frequency within the baroreflex band (typically 0.075–0.125 Hz; Lehrer, 2013) to maximize heart rate oscillations and baroreflex gain. However, given emerging evidence that the upper range of this band (0.10–0.14 Hz) may reflect stronger parasympathetic modulation (Pfurtscheller et al., 2018; Schneider & Schwerdtfeger, 2020), we subdivided the LF spectrum into lower, middle, and upper sub-bands (0.058–0.068, 0.075–0.093, 0.096–0.130 Hz) to test whether training-related gains in autonomic regulation and OFC plasticity were differentially associated with specific LF subcomponents, with particular emphasis on the upper-LF range.

An interesting question is whether the breathing-induced changes in OFC cortical volume that we previously demonstrated in our HRV-ER clinical trial (Yoo et al., 2022) might affect cognitive processes. Generally, patients with lesions in OFC show normal neuropsychological performance on cognitive tasks despite notable changes in behavior and emotion regulation and decision making (Angrilli et al., 1999; Eslinger & Damasio, 1985; Fujii et al., 2005; Namiki et al., 2008; Stuss et al., 1983). The specific OFC functions contributing to these behavior changes have been controversial. The somatic marker hypothesis posited that the OFC integrates body sensations to produce feelings that help guide decisions, whereas others subsequently argued that deficits were due to failures to inhibit no-longer appropriate responses. However, the notion that OFC-lesion-related behavioral deficits stem from failures of inhibitory control has also been disputed, with a newer view positing that the OFC encodes and stores new stimulus-reward associations (Rudebeck & Rich, 2018). The cognitive data collected in our HeartBEAM trial did not include decision or stimulus-reward measures, but did include cognitive tasks requiring inhibitory processing, such as the flanker tasks, which assesses the ability to suppress responses to irrelevant stimuli in the presence of competing information (Draheim et al., 2021). Thus, in our exploratory analyses examining whether the structural volume increases associated with heart rate oscillatory activity during practice sessions were associated with changes in cognition we hypothesized that we might see relationships with tasks involving inhibition, both in the set of neuropsychological tasks assessed pre and post intervention as well as in a set of brain training games (Lumosity; Steyvers & Schafer, 2020) that participants played daily during the intervention.

In addition, to better isolate relevant neural processes we were particularly interested in measures from separate sessions before and after the intervention in which participants were assessed using EEG while they completed an oddball task. We were interested in examining relationships between structural changes in the OFC and measures from the oddball task because the three-stimulus oddball task provides a measure of locus coeruleus (LC) activity (Aston-Jones et al., 2004; Dahl et al., 2025; Kuo et al., 2020; Polich, 2007), and the OFC and medial PFC project to the LC, particularly to its pericerulear dendritic zone (Jodo et al., 1998; Jodo & Aston-Jones, 1997), a region that modulates LC activity via local GABAergic interneurons (Aston-Jones et al., 2004; Kuo et al., 2020). The LC is the brain’s hub region for arousal-related noradrenergic activity and is also modulated by breathing-related signals, linking respiratory rhythms to central arousal regulation (Mather, 2020; Yackle et al., 2017).

We also assessed pupil dilation responses during the oddball task. In addition to electrophysiological measures, pupil dilation provides a sensitive and noninvasive index of LC–noradrenergic activity, with larger task-evoked pupil responses reflecting greater phasic LC engagement (Aston-Jones & Cohen, 2005; Murphy et al., 2011, 2014).

In the three-stimulus oddball task, participants are presented with a continuous sequence of standard stimuli punctuated by infrequent target stimuli (oddball) that require a response and infrequent salient distractors that do not require a response. This design engages two core processes: target detection and distractor discrimination, both essential for adaptive cognitive control. The oddball task reliably elicits the P300a and P300b components of the event-related potential (ERP), a positive deflection occurring approximately 300 ms after stimulus onset, reflecting the allocation of attentional resources and the updating of working memory representations, and has been linked to phasic activity of the LC–noradrenergic system (Nieuwenhuis et al., 2005; Polich, 2007; Vazey et al., 2018). Notably, the amplitude and latency of the P300 provide sensitive indices of cognitive processing efficiency and resource allocation (Kok, 2001). Furthermore, individual differences in autonomic regulation have been robustly linked to attentional control and cognitive performance, consistent with neurovisceral integration models that emphasize the role of cardiac vagal tone in prefrontal regulatory function (Park & Thayer, 2014; Thayer & Lane, 2009). Within this framework, heart rate variability has been shown to predict performance in P300 oddball–based tasks, providing empirical support for a link between cardiovascular dynamics and attentional processing efficiency (Kaufmann et al., 2012). In the present study, by examining both behavioral measures (reaction time) and neurophysiological markers (pupil dilation response, P300 amplitude and latency), we aimed to gain insight into how autonomic and cortical plasticity interact to modulate attentional control and cognitive flexibility.

In summary, to test whether heart rate oscillatory dynamics during training related to changes in OFC structure, we examined associations between HRV power in lower, middle, and upper LF sub-bands and post–pre changes in OFC volume, using both ROI-based and vertex-wise analyses. Partial least squares correlation (PLSC) analyses were then used to capture coordinated multivariate associations between OFC structural changes and oddball task performance measures. An overview of the study design and analytical framework linking training HRV, cortical plasticity, and cognitive performance is illustrated in Supplementary Figure S1.

## 2. Methods

### 2.1. Participants

We recruited 91 participants aged 50-70 years through the USC Healthy Minds subject pool, targeted mailings, community flyers, and the recruitment service Trialfacts between January and October 2023. Participants provided written informed consent approved by the University of Southern California (USC) Institutional Review Board. Eligibility criteria included: English fluency, post-menopausal status (for women), normal or corrected vision and hearing, body weight of at least 110 pounds, willingness to provide blood and urine samples, access to a home computer with a physical keyboard and reliable internet, possession of a mobile phone capable of receiving text messages, and a commitment to devote approximately 60 minutes per day for 12 weeks to the study and check an email account regularly. Exclusion criteria included any cardiac disease (e.g., arrhythmia, angina), respiratory disorders, neurological or psychiatric illness, diagnosed cognitive impairment, regular practice of breathing-based relaxation or meditation exceeding 1 hour per week, prior participation in HRV biofeedback studies in our lab, use of Lumosity in the past 6 months, inability to understand task instructions, and MRI contraindications. Cognitive functioning was assessed using the Montreal Cognitive Assessment (MoCA; Nasreddine et al., 2005). In the present dataset, the mean MoCA score was 26.4 (SD = 2.24; range = 21-30), within ranges seen in population-based samples (Rossetti et al., 2011). Participants had an average of 16.58 years of education (SD = 2.36; range = 12–24) and a mean age of 60.13 years (SD = 5.99; range = 50–70).

Of the 91 individuals who completed screening and provided written informed consent, 29 participants dropped out before being randomized to condition (most before or during the week of home assessments), 2 participants dropped out during the paced breathing intervention phase, and 5 participants did not complete one or more of the three MRI sessions (pre, mid, and post). The final analytic sample included 55 participants who completed all three MRI scans and the 9-week HRV biofeedback training, comprising 26 men and 29 women. There were no significant sex differences in age (*p* = .414), years of education (*p* = .810), or MoCA scores (*p* = .869).

### 2.2. Procedure

#### 2.2.1. Overview of the 12-week protocol schedule

Participants completed a 12-week protocol that included five laboratory visits and a 10-week home-based breathing and cognitive-training intervention. At baseline (Week 1), participants completed home assessments and questionnaires, followed by two laboratory visits (Week 2): (i) EEG and pupillometry during an auditory oddball task, and (ii) structural MRI (3T Siemens Prisma, 32-channel coil) and physiological baseline measurements (see Supplementary Fig. S2 for a visual summary).

Participants were then randomly assigned to one of two conditions:

● Slow-paced breathing condition: breathing cycles of 10, 12, or 15 s per breath.
● Random-paced breathing condition: breathing cycles of 4, 5, or 6 s per breath.

Both groups used identical lab-issued laptops and ear pulse sensors. Each day, participants completed two 15-min breathing sessions guided by a visual pacer and one set of Lumosity games. Breathing instructions differed by condition: slow-paced participants aimed to increase heart-rate oscillations (“relaxation score”), whereas random-paced participants aimed to reduce oscillations (“alertness score”).

A mid-intervention MRI (Week 7) assessed interim structural changes, and post-intervention sessions (Week 12) repeated all baseline procedures: EEG/pupillometry oddball task, cognitive testing, MRI, and biospecimen collection.

#### 2.2.2. Home assessments

During Week 1, participants completed initial home-based assessments consisting of cognitive tasks. The schedule varied by task: some measures were administered daily across six consecutive days, whereas others were administered only once or on a subset of days.

Thus, participants completed between one and six days of assessments, with a maximum duration of approximately 45 minutes per day (see Supplementary Table S1 for details). These home assessments were repeated in Week 12, after participants had completed all lab visits and the intervention period. For some measures, identical test forms were administered at both pre- and post-intervention time points, whereas for others, alternate forms were used to minimize practice effects.

#### 2.2.3. Lab visits

Participants visited the lab a total of five times: twice during Week 2 (pre-intervention), once at Week 7 (mid-intervention), and twice again in Week 12 (post-intervention). During the first lab visit, participants completed questionnaires assessing sleep quality and daily functioning (Jackson et al., 1990), as well as a battery of neuropsychological tests (Canning et al., 2004; Gollan et al., 2012; Tombaugh, 2004). They also performed an auditory oddball task while EEG and eye-tracking data were collected. During the second lab visit, urine and blood samples were collected, mood, stress, and sleep questionnaires were administered, and a baseline brain MRI scan was performed. At the end of this visit, participants were provided with the study laptop and ear sensor, which were used to complete two baseline pulse measurements during a 5-minute resting period, as well as a paced breathing exercise guided by a visual pacer set at 15 breaths per minute. The midpoint lab visit (Week 7) replicated the procedures from Visit 2, with the exception of the resting pulse measurement and paced breathing exercise. During Visit 4, participants repeated the oddball task with EEG and eye tracking and a battery of neuropsychological tests. Visit 5 (Week 12) mirrored Visit 2’s procedures, including the collection of biospecimens and questionnaires. Participants returned the study laptops and ear sensors at the end of this visit.

#### 2.2.4. Home practice

Participants were randomly assigned to one of two intervention conditions: slow-paced breathing or random-paced breathing. Both groups completed a 10-week home-based intervention using a lab-issued laptop and ear sensor.

During the first practice week (study Week 2), participants in both conditions played one set of *Lumosity* brain games daily (see Supplementary Table S2) and completed a 5-min resting pulse measurement each day. From Weeks 3–12, participants followed the same daily schedule with the addition of condition-specific breathing practice: they played one set of *Lumosity* games and completed two 15-min paced-breathing sessions per day, guided either at a slow-paced rhythm or at a random-paced rhythm depending on condition. The *Lumosity* battery sampled multiple cognitive domains, including memory, attention, mental flexibility, reasoning, processing speed, spatial orientation, and problem solving (Steyvers & Schafer, 2020). To balance content and minimize task-specific practice effects, the battery was divided into two sets of six games, which alternated across days. Details of the *Lumosity* games and their domain mappings are described in the Measures section (see Section 2.3.4, *Lumosity game*).

During the breathing exercises, heart rate was monitored using an earlobe sensor connected via USB to the study laptop. On the laptop screen, a visual pacer guided participants’ breathing, with a ball moving upward, downward, and horizontally along a line (See Supplementary Figure S3). Beneath the pacer, a real-time performance score based on participants’ heart rate oscillations was displayed, calculated in accordance with their assigned breathing condition. An audio cue was also provided as a guide. Before each breathing session, participants were instructed to sit upright in a chair with their feet flat on the floor and their hands resting in their laps. Each 15-minute breathing session was divided into three 5-minute segments (regimes). During each segment, the emWave Pro software (HeartMath Institute, 2020) continuously computed and displayed a real-time coherence score based on participants’ heart rate oscillations, with the displayed score reflecting performance according to the assigned breathing condition. For each segment, the coherence values generated in real time were averaged to produce a segment-level performance score. Based on these segment-level averages from previous sessions, a personalized breathing pace was assigned for each regime, ensuring that participants practiced breathing at a pace tailored to their individual performance (see Nashiro et al., 2024 for details).

##### Slow-paced breathing condition

Participants in this group followed slow breathing patterns of 10, 12, and 15 seconds per breath. A "relaxation score" reflecting heart rate oscillations in the 0.04–0.26 Hz frequency range was displayed during the exercise, with higher scores indicating greater oscillatory activity. Participants were encouraged to improve their relaxation scores through breathing focusing on the diaphragm: sitting with relaxed posture, loosening the neck and shoulders, placing one hand on the chest and the other on the abdomen, inhaling slowly through the nose while expanding the belly, and exhaling gently through the mouth as the belly contracted.

##### Random-paced breathing condition

Participants in this group were guided through breathing patterns of 4, 5, and 6 seconds per breath. An "alertness score" was provided during the exercise, where higher scores corresponded to reduced heart rate oscillations within the 0.04–0.26 Hz frequency range. To enhance performance, participants were instructed to breathe lightly and naturally, avoiding deep breaths.

#### 2.2.5. MRI scan parameters

The scans were conducted with a 3T Siemens MAGNETOM Prisma scanner with 32-channel head coil at the USC Dana and David Dornsife Neuroimaging Center. T1-weighted 3D structural MRI brain scans were acquired using a magnetization prepared rapid acquisition gradient echo (MPRAGE) sequence with repetition time (TR) = 2400 ms, echo time (TE) = 2.34 ms, slice thickness = 0.70 mm, flip angle = 8°, field of view = 224 mm, and voxel size = 0.7 x 0.7 x 0.7 mm3, with 256 slices collected (7:40 min). T2-weighted 3D structural MRI brain scans were acquired using a magnetization prepared rapid acquisition gradient echo (MPRAGE) sequence with TR = 3200 ms, TE = 566 ms, slice thickness = 0.70 mm, field of view = 224 mm, and voxel size = 0.7 x 0.7 x 0.7 mm3, with 256 slices collected (8:27 min).

#### 2.2.6. Oddball task setup

The oddball task sequence, design, and data analyses pipeline has previously been described in detail (Kim et al., 2026). In brief, stimuli were presented from a custom-built NZXT desktop computer (NZXT, Los Angeles, CA, USA) running MATLAB 2024b (Mathworks, Natick, MA, USA) with Psychophysics Toolbox extensions (Brainard, 1997). Visual stimuli were displayed on a Sun Microsystems 4472 CRT monitor (Oracle Corporation, Santa Clara, CA, USA) with a refresh rate of 85 Hz. Pupillometry data were acquired using the EyeLink 1000 Plus system (SR Research Ltd., Ottawa, Ontario, Canada) at a sampling rate of 1000 Hz. A 5-point calibration procedure was completed at the beginning of each run. and participants’ head position were stabilized using a chin rest from SR Research. Auditory stimuli were presented through inserted earphones designed for research (Model ER2; Etymotic Research, Inc., Fort Worth, TX, USA) with disposable foam ear tips (Models ER3-14B and ER3-14C; Etymotic Research).

All experimental procedures took place in a soundproof and electromagnetically shielded testing room constructed as a Faraday cage. Electrical equipment was powered externally and communication between the participant and experimenter was maintained via a two-way night-vision baby monitor system (Model SM935A; Kidsneed). EEG data were recorded using a 65-channel HydroCel Geodesic Sensor Net, sampled at 1000 Hz, and acquired via Net Station software (Version 5.4; Electrical Geodesics, Inc., Eugene, OR, USA). Channel Cz served as the online reference during data collection and data were re-referenced offline to the average of the mastoid electrodes during data pre-processing. Electrode impedance was continuously monitored throughout the session and maintained below 50 kΩ. Participants completed two runs of the oddball task. Each run consisted of 120 trials composed of 70% standard tones (500 Hz), 15% target tones (1000 Hz), and 15% distractor tones (salient auditory tone; burst of white noise), all presented at 75 dB. Inter-stimulus intervals (ISI) were jittered at 2.2, 2.3, or 2.4 seconds, and randomly distributed across trials equally often. Participants were required to make a motor response to all presented tones on a button box (SR Research): press the right button when hearing the target tone and press the left button when hearing all other tones.

### 2.3. Measures

#### 2.3.1. Cognitive measures

Cognitive functioning was assessed with a diverse set of tasks designed to measure multiple domains and administered online. Working memory was measured using an N-back task (Herrera et al., 2020), while episodic memory was assessed with both a face–name associative memory task (Polcher et al., 2017) and a pattern separation task (Wais et al., 2021). Executive function was examined with a task-switching task (Gajewski & Falkenstein, 2012; Olfers & Band, 2018) and a Flanker task (Draheim et al., 2021). Visuospatial function was measured with a spatial orientation task (Friedman et al., 2020), and verbal fluency was evaluated with the Thurstone Word Fluency Test (Spreen & Risser, 2003).

In addition to these online tasks, participants completed a brief battery of standard neuropsychological assessments during Visit 1and Visit 4, including the Trail Making Test Parts A and B (Tombaugh, 2004) and an Animal Fluency task (Canning et al., 2004).

#### 2.3.2. Lumosity games

Cognitive training during the intervention was implemented through 12 computerized brain training games on the Lumosity platform (https://www.lumosity.com/; Steyvers & Schafer, 2020). The games sampled six cognitive domains, including memory, attention, flexibility, reasoning, language, and mathematics. To reduce task-specific practice effects, the 12 games were divided into two sets of six, which alternated daily across the 10-week intervention period.

The memory domain included tasks that required recalling customers’ faces and food orders (*Familiar Faces*), identifying new items among visually similar ones (*Tidal Treasures*), and matching symbols across trials (*Memory Match*). Attention was targeted by tasks such as identifying the direction of a target bird amid distractors (*Lost in Migration*), evenly dividing visual stimuli without counting (*Splitting Seeds*), and navigating a maze that required spatial updating (*Penguin Pursuit*). Flexibility was assessed by shifting between competing stimulus–response rules, as in *Ebb and Flow*, *Brain Shift*, and *Color Match 2*. Reasoning was trained with tasks such as navigating a ship to a goal while avoiding obstacles (*Pirate Passage*). Language was assessed with a lexical fluency task in which participants generated words from letter stems (*Word Bubbles Rising*), and mathematical ability was trained with an arithmetic task in which participants solved problems before visual stimuli disappeared (*Raindrops*).

#### 2.3.3. Oddball pupil and p300 responses

The auditory oddball task (Kim et al., 2026) was used to assess attentional processing and cognitive control. Behavioral performance was indexed by accuracy and reaction time to target and non-target tones. Electrophysiological responses were quantified using the P300 component, a well-established marker of stimulus evaluation and attentional allocation (Polich, 2007). P300 amplitude and peak latency were extracted from the frontal midline electrode (Fz), given the relevance of frontal regions to cognitive control processes. Pupillometry was used to index autonomic and attentional engagement. Pupil dilation responses (PDRs) were calculated as the maximum stimulus-evoked change relative to pre-stimulus baseline (Mather et al., 2020).

### 2.4. Data processing

#### 2.4.1. MRI data processing

The images were processed following the HCP’s minimal preprocessing protocol (Glasser et al., 2013). Structural images underwent gradient distortion correction, were aligned to the anterior commissure-posterior commissure (AC-PC) axis, and had the brain extracted. T1-weighted and T2-weighted images were co-registered using a rigid body transformation enhanced by boundary-based registration (Greve & Fischl, 2009). Bias field correction was applied by taking the square root of the product of the T1-weighted and T2-weighted images after removing non-brain tissues (Glasser & Van Essen, 2011). Finally, the images were first linearly registered to MNI space using FSL FLIRT (12 degrees of freedom), followed by nonlinear registration using FSL FNIRT. The nonlinear warp was estimated using the MNI152 T1 2 mm template from the HCP templates directory, and final resampling was performed in the target MNI space resolution, as implemented in the HCP minimal preprocessing pipeline (post-v4.3.0).

FreeSurfer (v7.3.1) was employed for cortical parcellation and subcortical segmentation of the T1w volume, including total intracranial volume (TIV) estimation for head size correction (surfer.nmr.mgh.harvard.edu; Dale et al., 1999; Fischl et al., 2002). The FreeSurfer processing workflow included non-brain tissue removal, automated Talairach transformation, intensity normalization, segmentation, tessellation of the gray/white matter boundary, topology correction, and surface deformation. Additionally, T2w images were integrated into the FreeSurfer pipeline to refine the pial surface by distinguishing dura and vasculature.

After the initial FreeSurfer processing, FreeSurfer’s longitudinal processing pipeline was applied to enhance within-subject consistency across time points (Reuter et al., 2012). This approach involves creating an unbiased within-subject template using all available timepoints. Each timepoint is then reprocessed using this subject-specific template as an initial estimate for cortical reconstruction and segmentation, thereby reducing segmentation variability and increasing statistical power in detecting cortical and subcortical changes over time.

Finally, using a parcellation algorithm, the individual brains were mapped to the Desikan–Killiany probabilistic cortical atlas based on anatomic landmarks and cortical geometry (Desikan et al., 2006). The cortical volumes of individual participants were then extracted directly from FreeSurfer. Cortical volumes for bilateral lateral and medial OFC were used in the atlas-based ROI analysis.

All images were visually inspected at each processing stage and excluded if they exhibited significant quality issues or prominent artifacts. Cortical quality control was performed using the ENIGMA Quality Control Protocol 2.0, which involves visual inspection of cortical segmentations for each scan (Thompson et al., 2020). No scans in the present study showed significant quality problems or artifacts, and therefore no participants were excluded on the basis of imaging quality.

#### 2.4.2. Heart rate variability during home training

Participants’ pulse was measured using the emWave Pro software (HeartMath Institute, 2020) in combination with an infrared pulse plethysmograph (PPG) ear sensor. The pulse waveform was recorded at a sampling rate of 370 Hz. Inter-beat interval (IBI) data were extracted after the removal of ectopic beats and other artifacts, accomplished through a built-in correction process within the emWave Pro software. The extracted IBI data were segmented into three 5-minute regimes within each 15-minute session, with breathing pace individually adjusted for each regime. To account for transitions between regimes, the first and last 15 seconds of each regime were excluded, resulting in a maximum of 4 minutes and 30 seconds of data analyzed per regime. In cases where noise or signal loss led to shorter data segments, only those regimes with a minimum of 3 minutes and 30 seconds of usable data were included in the analysis.

Heart rate variability (HRV) was processed from the IBI data corresponding to each regime using the RHRV package in R (Rodríguez-Liñares et al., 2011). The data were pre-processed to remove ectopic beats and artifacts, and then interpolated at 4 Hz to produce continuous heart rate data (García Martínez et al., 2024).

Time-domain analysis involved calculating the root mean square of successive differences between adjacent RR intervals (RMSSD) and the standard deviation of all normal RR intervals (SDNN) over 60-second segments. Frequency-domain analysis included estimating power spectral density (PSD) and calculating spectral power for standard and custom frequency bands. The standard frequency bands included low frequency (LF; 0.04–0.15 Hz), high frequency (HF; 0.15–0.4 Hz), and total power. In addition to these standard bands, custom frequency bands were defined to capture specific subcomponents of the LF range, including upper low frequency (0.096–0.13 Hz), middle low frequency (0.075–0.093 Hz), and lower low frequency (0.058–0.068 Hz). These custom bands were established to subdivide the baroreflex frequency range more precisely and to explore whether specific frequency components within paced breathing-based HRV biofeedback training are associated with greater structural brain changes. For each regime, the calculated time-domain and frequency-domain HRV metrics were averaged across all sessions during the intervention period. These averaged values were used in subsequent statistical analyses to represent each participant’s overall performance throughout the training.

The Shapiro–Wilk test was run for heart rate and HRV metrics to ensure normal distribution. All these metrics, except mean HR, were not normally distributed (p < 0.05). To correct for this, SDNN, RMSSD, HF power, LF power, total power, and sub-band power within the baroreflex frequency range were transformed using the natural log function.

#### 2.4.3. Oddball data processing

EEG data were processed using the standardized and semi-automated MADE pipeline (Debnath et al., 2020; Leach et al., 2020), previously described in detail (Kim et al., 2026). Raw continuous EEG data were first exported to MATLAB for offline preprocessing using the EEGLAB toolbox (Delorme & Makeig, 2004). Data were down sampled to 500 Hz, high-pass filtered at 0.1 Hz, and low-pass filtered at 50 Hz. Artifact-laden channels were identified and removed using the FASTER algorithm (Nolan et al., 2010). Next, independent component analysis (ICA) was performed by first creating a copy of the dataset. This copy was high-pass filtered at 1 Hz and segmented into 1-second epochs. Noisy segments and EMG artifacts were excluded using a voltage threshold of ±1000 *mV* and spectral threshold (range -100 dB to +30 dB) within the 20-40 Hz frequency band. Channels exhibiting artifacts in more than 20% of epochs were excluded from both the original and the copied dataset. ICA was then performed on the filtered copy and the resulting ICA weights were transferred back to the original dataset (Viola et al., 2010). Artifactual components were then automatically identified and removed using the Adjusted-ADJUST algorithm (Leach et al., 2020). The cleaned data were segmented into 1500 ms epochs, beginning 500 ms before stimulus onset. To further address residual artifacts, we applied a two-step rejection procedure (Morales et al., 2022). First, epochs were discarded if voltages from ocular electrodes (E1, E2, E5, E10, E11, and E17) exceeded ±125 *mV* to remove residual ocular activity. Second, for non-ocular channels, values exceeding ±150 *mV* were interpolated at the epoch level. If more than 10% of channels (excluding previously removed global channels) exceeded ±125 in an epoch, the entire epoch was rejected. Missing channels were interpolated using spherical spline interpolation and data were re-referenced to the average of the mastoid electrodes prior to quantification.

P300 amplitude and peak latency were automatically extracted from stimulus-evoked ERP waveforms using custom MATLAB scripts using the EEGLAB toolbox (Delorme & Makeig, 2004). Baseline correction was applied using the -200 to 0 ms pre-stimulus window and ERPs were averaged across all artifact-free trials. Given the location of the orbitofrontal cortex and the maximal P300 in the midline (Polich, 2007), we extracted P300 components from the Fz electrode. To define the temporal window of the P300 component, a data-driven approach was used. Using active - passive task difference waveforms data from the larger experiment (Kim et al., 2026), we identified that participants’ P300 components had a 288-432 post-stimulus window. P300 amplitudes were quantified by computing the area under the curve (AUC) within the respective time windows for each group. Peak latencies were determined using MATLAB’s *findpeaks* function to identify the maximum positive peak within the defined window, with the latency corresponding to the timing of that peak. Given our interest in the OFC, we used values from the frontal Fz electrode.

Eye-tracking data were stored in EDF format and were imported into MATLAB using the *edf2mex* MEX program. Pupil data were preprocessed using the automated artifact removal algorithm *ET-remove-artifacts* (Mather et al., 2020), which identifies blink-related artifacts by detecting abrupt changes in pupil velocity and linearly interpolates across the start and end points of each artifact to generate a cleaned time series. Pupil dilation responses (PDRs) were quantified as the maximum change in pupil size relative to a 500 ms pre-stimulus baseline window. The PDR was defined as the maximal pupil dilation occurring within the 2000 ms window following stimulus onset.

#### 2.4.4. Cognitive task data processing

In the N-back task, accuracy was defined as the proportion of correct responses and mean RTs were calculated for correct trials; both measures were computed separately for the 0-back, 1-back, and 2-back conditions. In the face–name associative memory task, accuracy was measured as the proportion of correctly identified face–name pairs. For the pattern separation task, accuracy was calculated as the proportion of correct rejections to lure items and the proportion of false alarms to novel items.

Executive function tasks were also processed in the same way. For the task-switching task, accuracy and mean RTs were computed separately for each condition. Specifically, hit rates and mean RTs were calculated for single-task and mixed-task trials, as well as for switch and nonswitch trials within the mixed-task blocks. In the response-deadline version of the

Flanker task, only accuracy for congruent and incongruent trials was analyzed, as RTs are not reliable indicators of attentional control in this paradigm (Draheim et al., 2021). For the spatial orientation task, performance was scored based on the angular difference between the correct orientation and the participant’s response, with smaller values indicating higher accuracy. This error-based metric was retained to preserve the original task scoring. In the verbal fluency task, participants were asked to generate as many unique correct words as possible within a limited time period, and performance was scored as the total number of words produced. Reaction times were not analyzed.

Standard neuropsychological measures were processed using conventional scoring procedures. For the Trail Making Test Parts A and B and the Animal Fluency task, normed T-scores were used as performance indices, with higher scores indicating better performance. These T-scores were normed based on sex and years of education, using Caucasian normative data for all participants. For all neuropsychological measures included in the analyses, higher scores reflected better performance.

#### 2.4.5. Lumosity game data processing

Participants played 12 brain training games from six cognitive domains (attention, flexibility, language, math, memory, reasoning) on the Lumosity platform. The games were divided into two alternating daily sets, and across the 10-week intervention participants completed each game approximately 32–34 times.

Analyses focused on raw performance scores, which were standardized within each game across participants and game plays. To account for gradual learning curves and missing data, performance scores for each game were aggregated across predefined bins of game plays. These bins reflected equivalent numbers of game plays distributed across the 10-week intervention period. Pre- and post-intervention performance were defined as the mean scores from the first and last game-play bins, respectively, calculated separately for each game.

### 2.5. Statistical analyses

#### 2.5.1. Partial correlation analysis of training-related HRV and OFC volume

To investigate the relationship between heart rate oscillatory power in the baroreflex frequency range during training and changes in OFC volume, we conducted a series of partial correlation analysis. Using the Freesurfer atlas-based OFC segmentation, we extracted OFC volumes at each timepoint and controlled for pre-intervention volume values. Specifically, we examined the partial correlation between post-intervention OFC volume and major heart rate variability (HRV) indices as well as power values within the baroreflex frequency range during training. All partial correlation coefficients were reported along with significance corrected for multiple comparisons using the Benjamini-Hochberg method (Benjamini & Hochberg, 1995).

To evaluate whether outliers might influence the partial correlation results, we first checked for outliers in the raw HRV indices and bilateral OFC volumes using standard univariate criteria (e.g., values exceeding 3 × IQR from the quartiles). No observations were flagged through this procedure; therefore, Pearson correlations among the global HRV indices and bilateral OFC volumes were computed and reported using the full dataset. While visualizing the partial correlations for our variables of interest, including the left OFC volumes and the three low-frequency power subcomponents, we observed that the residual values indicated the presence of potential bivariate outliers. Specifically, after residualizing post-intervention OFC volume for pre-intervention values, visual inspection of the standardized residual scatterplots indicated the presence of observations that appeared extreme within the joint residual distribution, despite not being detectable at the univariate level.

To address the potential influence of these residual-based leverage points, we employed two complementary robust correlation approaches when computing and visualizing the partial correlations for the left OFC and training-related LF power measures. Spearman’s rank correlation (ρ) provided a rank-based estimate that reduces sensitivity to extreme values without excluding data points, whereas Shepherd’s pi (π) correlation (Schwarzkopf et al., 2012) explicitly identifies and removes bivariate outliers using L1 (Manhattan) distance from the bivariate median in the residual space. By reporting both methods, we were able to transparently assess the robustness and stability of the observed partial correlations under alternative assumptions about outlier influence.

#### 2.5.2. Whole-brain vertex-wise analysis of cortical volume

Changes in subcortical volume and cortical thickness across three timepoints during HRV biofeedback training were assessed using the symmetrized percentage change (SPC), which represents the annualized rate of change (in mm³/year for volume) relative to the mean volume across all three timepoints. Cortical data were prepared by applying smoothing (10 mm full width at half maximum) and mapping the images to the Freesurfer average subject template using the long_mris_slopes command, which also computes SPC. To examine whether cortical volume changes were associated with log-transformed power values from each frequency band during training, a general linear model (GLM) analysis was conducted using mri_glmfit with log power value as a predictor and intracranial volume (ICV) included as a nuisance variable.

Correction for multiple comparisons was performed using a cluster-wise Monte Carlo Z simulation (1000 iterations), with an absolute cluster-forming threshold of *p* < 0.005, a cluster-wise threshold of *p* < 0.05. Significant clusters were visualized using Freeview.

#### 2.5.3. Partial correlations between the left OFC volume and power within a broad frequency range covering all breathing paces

Partial correlation analysis was conducted between the power values within the frequency range covering both random breathing and resonance breathing conditions (going from 4 to 20 seconds per breath) and the OFC volume. Power spectral density (PSD) values were extracted at breathing-related frequencies corresponding to 4-20 second breathing cycles. Specifically, power values were computed at 1-second intervals for breathing periods of 4, 5, 6, … up to 20 seconds per breath (i.e., 1/4, 1/5, …, 1/20 Hz, equivalent to 0.25, 0.20, …, 0.05 Hz). For each target frequency, we employed a nearest-neighbor approach to identify the frequency bin with the minimum absolute distance from the target frequency in the continuous PSD spectrum. The power value associated with this nearest frequency bin was then extracted as the representative power at the target frequency. To assess the relationship between these log-transformed power values and changes in ROIs (clusters in the left hemisphere lateral and medial OFC identified from the whole-brain analysis that were correlated with the upper low-frequency range), partial correlations were calculated between log power and post-intervention OFC ROI volume, while controlling for pre-intervention volume. The correlation values for each frequency were then compared to investigate how the relationships vary across this broad range of breathing frequencies. All partial correlation coefficients were reported along with significance corrected for multiple comparisons using the Benjamini-Hochberg method (Benjamini & Hochberg, 1995).

#### 2.5.4. Vertexwise partial least squares correlation (PLSC) analysis of OFC cluster volume changes and behavioral performance

Vertexwise PLSC was conducted to investigate multivariate associations between changes in brain structure and changes in behavioral performance following the intervention (Krishnan et al., 2011). This method is particularly advantageous for neuroimaging–behavior analyses because it is robust to multicollinearity among predictors, a common limitation in ROI-based datasets, and can capture latent dimensions that link distributed neural changes with complex behavioral profiles. Prior to running PLSC, we performed multivariate outlier screening using Mahalanobis distance on the set of behavioral variables, after z-scoring all measures. Outliers were defined relative to the chi-square distribution with degrees of freedom equal to the number of variables (α = .01). This procedure flagged one participant in the Lumosity dataset. This case was excluded from the corresponding analyses, and we confirmed through sensitivity checks that the significance of the extracted latent variables and the bootstrap stability patterns were unchanged. This suggests that the reported results are robust to the inclusion or exclusion of these extreme observations.

For the vertexwise PLSC, regions of interest were defined as two OFC clusters in the left lateral OFC (LOFC) and medial OFC (MOFC) that showed significant associations with upper LF power during training in the preceding GLM analysis. The X matrix comprised post–pre difference scores of vertexwise volume estimates across all vertices within each of the two clusters. On the Y side, separate PLSC analyses were applied to three behavioral domains: 1) oddball task performance, 2) cognitive and neuropsychological test measures, and 3) gamified Lumosity tasks. For the OFC–oddball PLSC, the Y matrix comprised post–pre difference scores for eight oddball measures: reaction time (target, distractor), pupil-dilation response (target, distractor), P300 amplitude (target, distractor), and P300 latency (target, distractor). For the OFC–cognitive-task PLSC, the Y matrix contained post–pre difference scores from a broad task battery assessing working memory, executive control, episodic memory, and language. Working memory was indexed by N-back performance, including hit rates and reaction times (RTs) for 0-, 1-, and 2-back conditions. Executive control was assessed with task switching (hit rates and RTs for single, mixed, switch, and nonswitch trials) and flanker interference (accuracy for congruent vs. incongruent trials). Episodic memory was measured with the pattern separation index (lure correct rejections – novel false alarms) and the CVLT- learning sum, short-delay free recall, and long-delay free recall. Associative memory was indexed by accuracy on a face–name task. Language abilities were assessed with a letter fluency task. In addition to these cognitive tasks, three standard neuropsychological test scores were also included: normed scores from Trails A and Trails B, and performance on an animal fluency task. For the OFC–Lumosity game PLSC, the Y matrix comprised post–pre difference scores from 12 web-based games (standardized game performance metrics): Lost in Migration, Ebb and Flow, Penguin Pursuit, Color Match, Memory Match, Tidal Treasures, Familiar Faces, Pirate Passage, Brain Shift, Splitting Seeds, Word Bubbles, and Raindrops. All behavioral variables were expressed as post–pre differences and z-scored across participants before entry into PLSC.

PLSC identifies latent variables (LVs) that capture the maximal shared covariance between the two datasets (Krishnan et al., 2011). To assess the statistical significance of the extracted latent variable(s), permutation testing (n = 5000) was conducted by randomly shuffling the rows of the Y matrix and recomputing the singular values to generate a null distribution. The *p*-value was determined as the proportion of permuted singular values that exceeded the original singular value. To evaluate the reliability of the saliences (weights) of the individual ROIs and behavioral measures, bootstrap resampling (n = 5000) with replacement was performed, resulting in bootstrap ratios (salience divided by its standard error). Variables with |BSR| > 1.96 were considered statistically significant. This approach allowed us to capture coordinated brain–behavior associations that may not be apparent when variables are examined individually (Krishnan et al., 2011).

## 3. Results

### 3.1. HRV during training and volume changes in OFC

Details about the baseline characteristics of the participants can be found in Supplementary Table S3. To examine the relationship between HRV during training and changes in OFC volume based on the Freesurfer Desikan–Killiany atlas (Fig 1), a partial correlation analysis was conducted using post-intervention volume while controlling for pre-intervention volume. As shown in Table 1, several HRV indices showed significant positive correlations with volume in the left medial OFC, including log-transformed RMSSD (*r* = .37, *p* = .006), SDNN (*r* = .36, *p* = .008), total power (*r* = .34, *p* = .013), HF power (*r* = .40, *p* = .003), LF power (*r* = .29, *p* = .033), baroreflex power (0.075–0.130 Hz; *r* = .36, *p* = .007), middle LF power (0.075–0.093 Hz; *r* = .30, *p* = .029) and upper LF power (0.096–0.130 Hz; *r* = .43, *p* = .001). In the left lateral OFC, HF power (*r* = .29, *p* = .036) and upper low-frequency power (*r* = .32, *p* = .019) were also significantly associated with post-intervention volume. After applying the Benjamini–Hochberg false discovery rate (FDR) correction, all associations in the left medial OFC remained significant except for log lower LF power. In contrast, none of the associations in the left lateral OFC remained significant after FDR correction. In contrast, no significant associations were observed in the right OFC, even at uncorrected thresholds (see Table 1).

**Figure 1.**
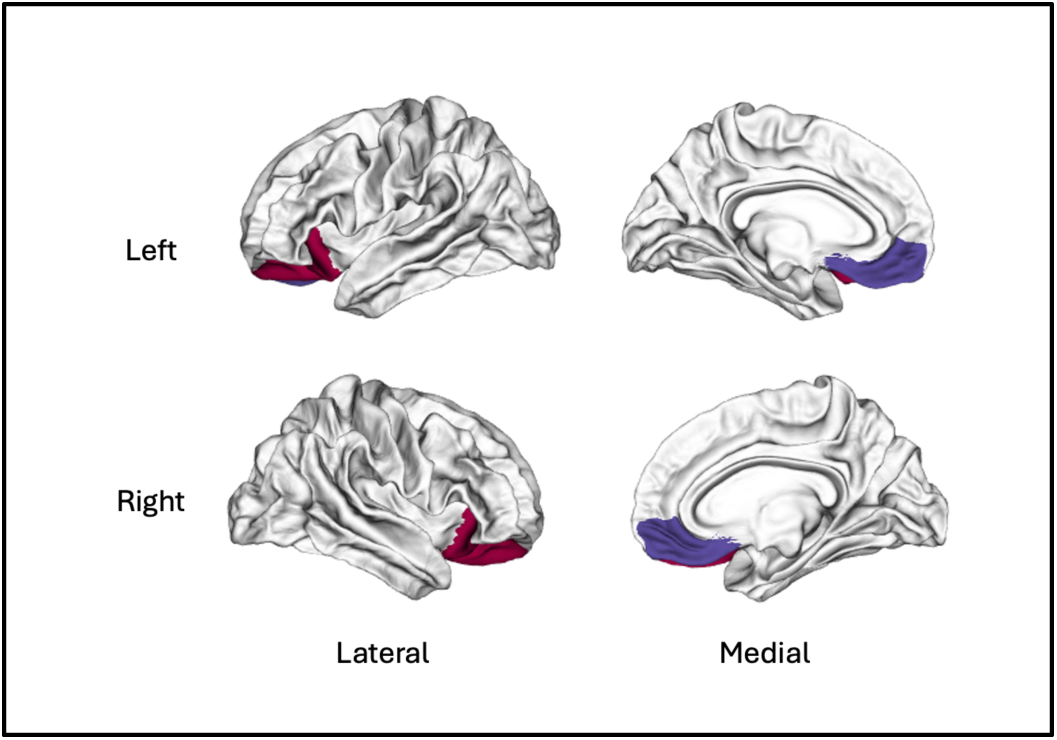
Orbitofrontal cortex (OFC) subregions defined using the Freesurfer Desikan–Killiany atlas. *Note.* Colored regions indicate lateral (red) and medial (blue) OFC subdivisions in the left and right hemispheres. Surface renderings are presented in lateral (left column) and medial (right column) views. These regions of interest were used to extract cortical volume measures for subsequent partial correlation analyses.

**Table 1.** Partial correlations between HRV during training and OFC volume at post-intervention, controlling for volume at pre-intervention

|  |  |  | Log<br>RMSSD | Log<br>SDNN | Log<br>total<br>power | Log<br>HF<br>power | Log<br>LF<br>power | Log<br>baroreflex<br>power | Log<br>lower<br>LFP | Log<br>middle<br>LFP | Log<br>upper<br>LFP |
| --- | --- | --- | --- | --- | --- | --- | --- | --- | --- | --- | --- |
| Left | Lateral<br>OFC | *r* | .235 | .196 | .155 | .286* | .094 | .231 | .069 | .188 | .318* |
|  |  | *p* | .087 | .156 | .264 | .036 | .499 | .092 | .619 | .175 | .019 |
|  | Medial<br>OFC | *r* | **.369\*\*** | **.357\*\*** | **.337\*** | **.401\*\*** | **.290\*** | **.364\*\*** | .157 | **.297\*** | **.425\*\*** |
|  |  | *p* | .006 | .008 | .013 | .003 | .033 | .007 | .258 | .029 | .001 |
| Right | Lateral<br>OFC | *r* | .151 | .107 | .038 | .189 | -.036 | .08 | -.035 | .051 | .165 |
|  |  | *p* | .277 | .440 | .787 | .171 | .794 | .566 | .804 | .714 | .232 |
|  | Medial<br>OFC | *r* | .132 | .082 | .033 | .160 | -.026 | .093 | -.082 | .033 | .208 |
|  |  | *p* | .341 | .556 | .814 | .248 | .854 | .502 | .556 | .815 | .131 |

*Note.* \* $p < .05$ ; \*\* $p < .01$ , \*\*\* $p < .001$ , 2-tailed. Correlations were controlled for volume at pre-intervention. Values in bold indicate statistical significance after FDR correction using the Benjamini-Hochberg method. RMSSD = root mean square of successive differences; SDNN = standard deviation of NN intervals; LFP = low frequency power. Frequency bands were defined as follows: baroreflex power = 0.075–0.130 Hz; lower LFP = 0.058–0.068 Hz; middle LFP = 0.075–0.093 Hz; upper LFP = 0.096–0.130 Hz.

Figure 2 presents partial correlations between log-transformed low-frequency power during training and post-intervention left OFC volume (controlling for pre-intervention volume), separately for lower (0.058–0.068 Hz), middle (0.075–0.093 Hz), and upper (0.096–0.130 Hz) frequency sub-bands. To address potential bivariate outliers observed in the residual space, we report both Spearman’s rank correlation (*ρ*; all data included) and Shepherd’s π correlation (outliers removed). In the left lateral OFC (Panels A–C), upper low-frequency power showed significant positive associations (*ρ* = .30, *p* = .028; *π* = .27, *p* = .050), and middle low-frequency power became significant after outlier removal (*π* = .30, *p* = .028). In the left medial OFC (Panels D–F), upper low-frequency power demonstrated the strongest associations (*ρ* = .38, *p* = .004; *π* = .35, *p* = .010), while middle low-frequency power showed significance only with Spearman’s correlation (*ρ* = .27, *p* = .043). Lower low-frequency power showed no significant relationships in either region. This pattern suggests a frequency-dependent gradient in both lateral and medial OFC, with the upper baroreflex sub-band (0.096–0.130 Hz) showing the most robust relationships with OFC volume. The convergence of results between rank-based and outlier-removal approaches indicates that these associations are stable and not driven by extreme observations.

**Figure 2.**
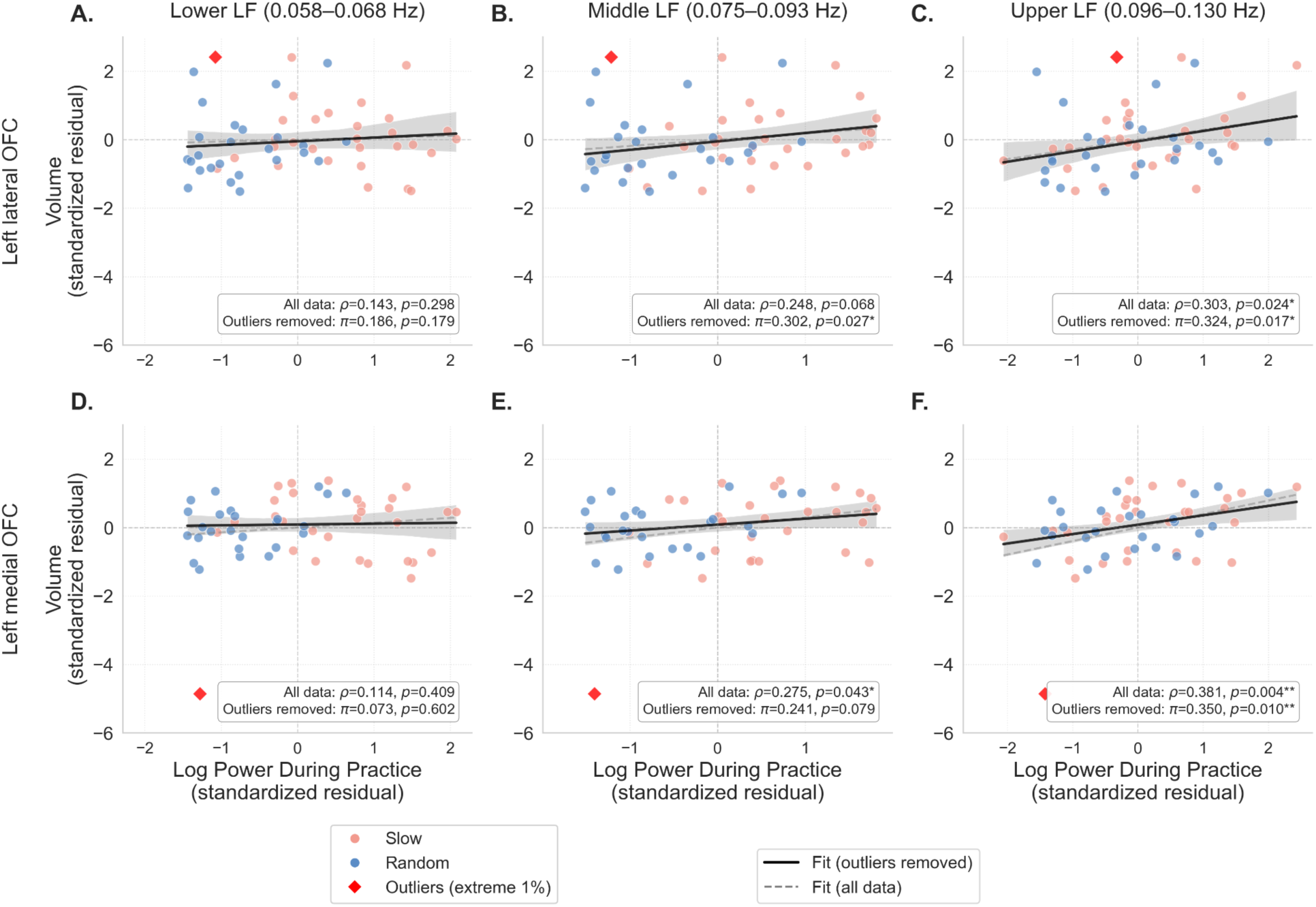
Robust partial correlations between log-transformed low-frequency power during training and post-intervention left OFC volume (controlling for pre-intervention left OFC volume) across three sub-bands. *Note*. Frequency bands were defined as follows: lower (0.058–0.068 Hz), middle (0.075–0.093 Hz), and upper (0.096–0.130 Hz). Panels A–C display the left lateral OFC; panels D–F display the left medial OFC. Pre-intervention volume was controlled for in all analyses. Slow = slow-paced breathing condition; Random = random-paced breathing condition. Data points are colored by training condition (Slow in coral, Random in blue), with bivariate outliers identified using the 99th percentile threshold marked in red. Gray dashed lines represent Spearman’s rank correlation (ρ) using all data; black solid lines represent Shepherd’s π correlation after removing bivariate outliers, with shaded regions indicating 95% confidence intervals.

### 3.2. Whole brain analysis: Cortical volume clusters associated with HRV low-frequency power

To further examine spatial patterns of brain regions associated with HRV power across the low-frequency (LF) range, surface-based whole-brain analyses were conducted using FreeSurfer’s general linear model (mri_glmfit).

Figures 3A-C illustrate significant clusters showing associations between log-transformed power within LF sub-bands and cortical volume changes across pre-, mid-, and post-intervention scans. Cluster-wise statistical details, including peak MNI coordinates, cluster sizes, and corrected *p*-values, are summarized in Table 1. Significant positive associations were observed in the upper low-frequency range (0.096–0.130 Hz), particularly in the left lateral and medial orbitofrontal cortex (OFC). These clusters showed the most robust effects among all frequency bands, with cluster-wise corrected *p*-values (CWP) of .001 for both regions. These findings are visualized in Figure 3A. In the middle low-frequency range (0.075–0.093 Hz), a more spatially restricted but still significant cluster was found in the left lateral OFC (CWP = .030), indicating a consistent association of changes in this region with HRV power during practice sessions. This result is shown in Figure 3B.

**Figure 3.**
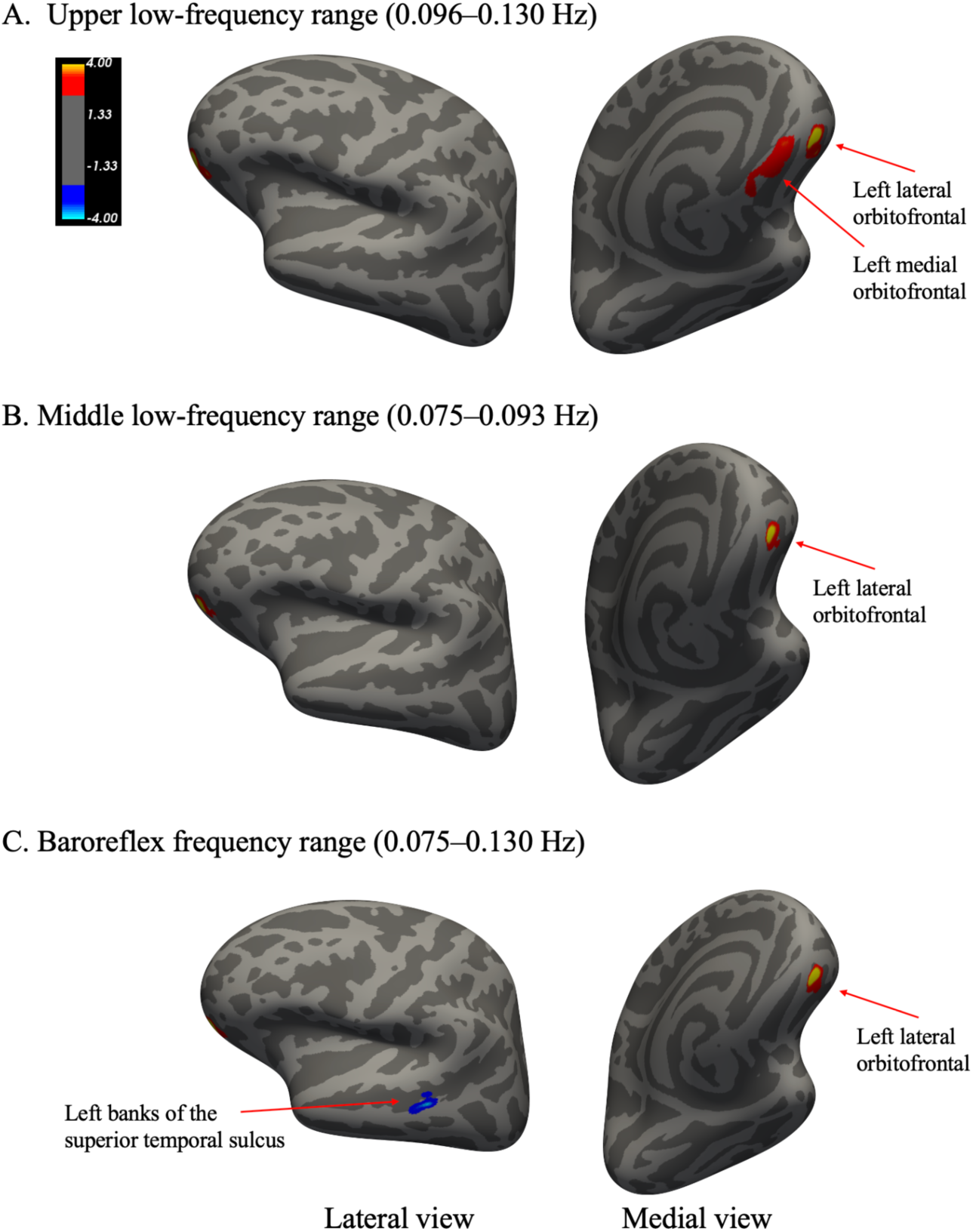
Significant cortical clusters associated with cortical volume change and log power during training in three low-frequency bands: (A) upper low-frequency power (0.096–0.130 Hz), (B) middle low-frequency power (0.075–0.093 Hz), and (C) baroreflex frequency range (0.075–0.130 Hz) *Note.* Colored regions indicate direction of association: red–yellow shows positive associations, and blue indicates negative associations. Results were corrected using Monte Carlo Z simulation (cluster forming threshold: p < .005; cluster-wise threshold: p < .05). No significant clusters were found in the lower low-frequency range (0.058–0.068 Hz) or in the right hemisphere.

When the middle and upper low-frequency bands were combined to form the baroreflex frequency range (0.075–0.130 Hz), the resulting pattern was similar. A significant cluster again appeared in the left lateral OFC (CWP = .006), and an additional small but significant negative cluster was observed in the left banks of the superior temporal sulcus (bankssts; CWP = .022), suggesting region-specific inverse associations outside the prefrontal cortex. These findings are illustrated in Figure 3C. In contrast, no significant clusters were found for the lower low-frequency range (0.058–0.068 Hz). Moreover, no significant clusters emerged in the right hemisphere for any frequency band.

**Table 2.**
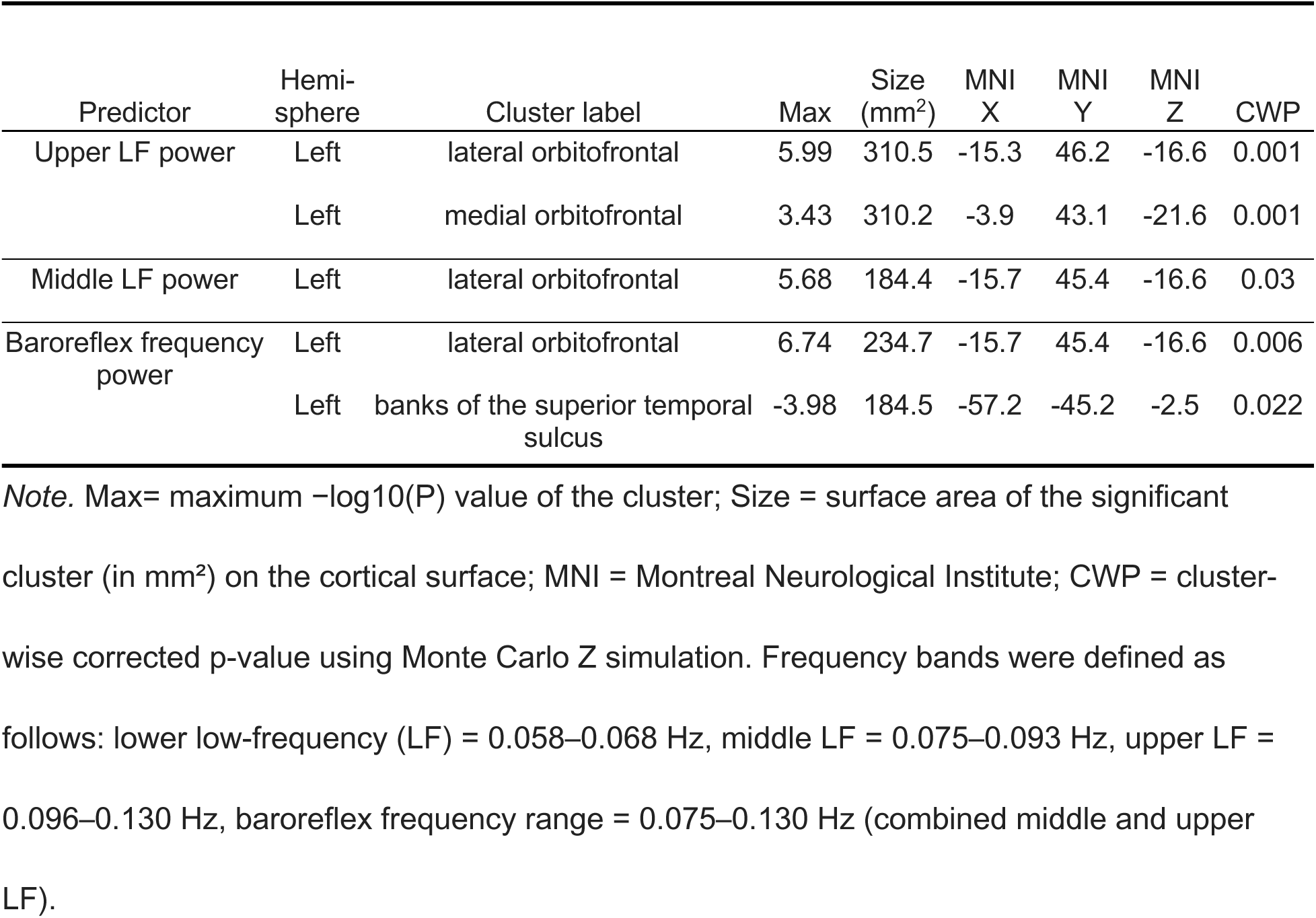
Significant Clusters Showing Associations Between Log Power During Training and Cortical Volume Changes

| Predictor | Hemi-sphere | Cluster label | Max | Size (mm<sup>2</sup>) | MNI X | MNI Y | MNI Z | CWP |
| --- | --- | --- | --- | --- | --- | --- | --- | --- |
| Upper LF power | Left | lateral orbitofrontal | 5.99 | 310.5 | -15.3 | 46.2 | -16.6 | 0.001 |
|  | Left | medial orbitofrontal | 3.43 | 310.2 | -3.9 | 43.1 | -21.6 | 0.001 |
| Middle LF power | Left | lateral orbitofrontal | 5.68 | 184.4 | -15.7 | 45.4 | -16.6 | 0.03 |
| Baroreflex frequency power | Left | lateral orbitofrontal | 6.74 | 234.7 | -15.7 | 45.4 | -16.6 | 0.006 |
|  | Left | banks of the superior temporal sulcus | -3.98 | 184.5 | -57.2 | -45.2 | -2.5 | 0.022 |

*Note.* Max= maximum $-\log_{10}(P)$ value of the cluster; Size = surface area of the significant cluster (in mm<sup>2</sup>) on the cortical surface; MNI = Montreal Neurological Institute; CWP = cluster-wise corrected p-value using Monte Carlo Z simulation. Frequency bands were defined as follows: lower low-frequency (LF) = 0.058–0.068 Hz, middle LF = 0.075–0.093 Hz, upper LF = 0.096–0.130 Hz, baroreflex frequency range = 0.075–0.130 Hz (combined middle and upper LF).

### 3.3. Partial correlations between OFC cluster volume change and HRV power within the breathing frequency range

To examine frequency-specific associations between heart rate oscillations and training-related brain changes, we conducted partial correlations between post-intervention OFC cluster volumes (identified in Figure 3A) and spectral power derived from heart rate data collected during training. To examine frequency-specific associations, we calculated spectral power at discrete frequencies corresponding to breathing cycles ranging from 4 to 20 seconds (i.e., 0.05–0.25 Hz). This frequency range was selected to broadly encompass the range of respiration rates observed across natural and paced breathing, allowing comparison of HRV power across the full spectrum of physiologically relevant breathing rhythms. OFC volume at pre-intervention was included as a covariate to control for baseline differences.

As shown in Figure 4, the strongest associations were observed around 0.083 Hz, a frequency situated near the transition point between the middle and upper low-frequency bands. At this frequency, correlation coefficients peaked at *r* = .50 in the lateral OFC and *r* = .49 in the medial OFC cluster. Importantly, correlations remained relatively strong across much of the upper low-frequency range, suggesting that power within this band during home practice is consistently associated with OFC volume change.

**Figure 4.**
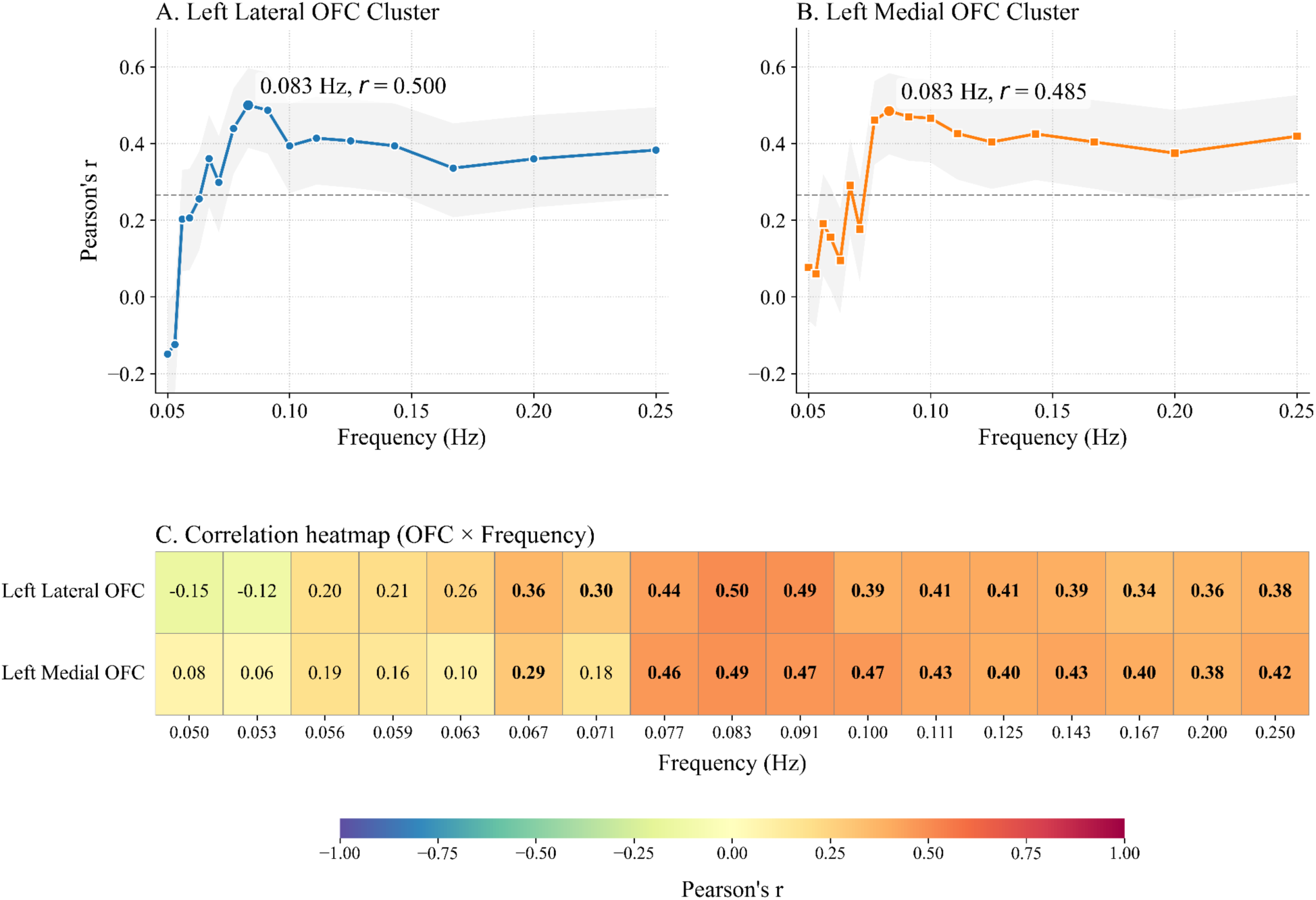
Partial Correlations Between Power at Each Frequency (0.05–0.25 Hz) and Post-Intervention OFC Cluster Volume, Controlling for Baseline Volume. *Note*. Panels A–B show partial correlations (r) between log power at each frequency and post-intervention OFC cluster volume, controlling for baseline (pre-intervention) volume. A = left lateral OFC cluster; B = left medial OFC cluster. Shaded bands denote ±1 standard error (SE) around r, and the gray dashed line marks the two-tailed *p* = .05 correlation threshold. The peak frequency and its r are annotated on each curve. Panel C summarizes the partial r values across frequencies for both clusters as a heatmap; warmer colors indicate stronger positive associations, and the numeric values inside each cell are r (boldface indicates values surviving FDR correction at *p* < .05, Benjamini–Hochberg method).

In contrast, associations in the lower low-frequency range (approximately 0.058–0.068 Hz) mostly did not reach significance, with correlation coefficients generally below .20 across both clusters.

### 3.4. Brain–behavior associations between OFC volume changes and oddball task measure changes

In the whole-brain GLM analysis, we found that upper LF power obtained during training was positively associated with structural volume changes in two clusters located in the left lateral and medial OFC. To further investigate whether these OFC structural changes were linked to changes in oddball task performance, we used the identified clusters as regions of interest and conducted a voxelwise partial least squares correlation (PLSC) analysis with post–pre difference scores. This analysis identified two significant latent variables (LV1, permutation *p* = .002; LV2, permutation *p* = .039), each capturing reliable associations between OFC volume changes and task-evoked behavioral outcomes (Figure 5).

**Figure 5.**
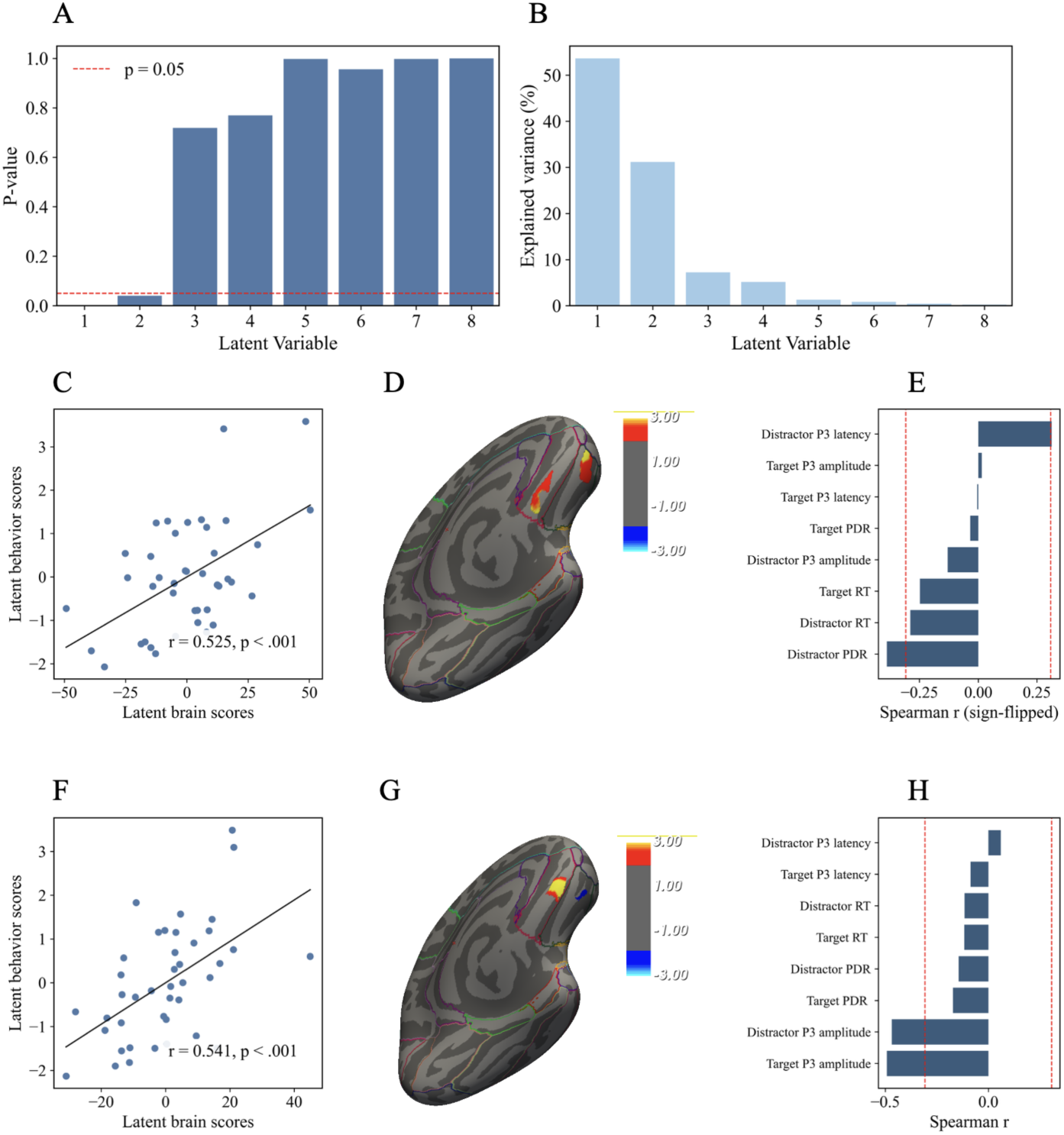
Vertexwise partial least squares correlation (PLSC) results: OFC volume change associated with oddball task performance change. *Note*. Panels A–B show significance testing of latent variables. Panel A displays permutation *P*-values of each latent variable (LV). Panel B shows the explained variance (%) for each LV. Panels C–E present results for LV1. Panel C shows the correlation between latent brain and behavior scores. Panel D illustrates OFC clusters that contributed significantly to LV1, with bootstrap ratios thresholded at ±1.96. Panel E shows Spearman correlations between oddball behavioral measures and LV1 brain scores. Panels F–H present results for LV2. Panel F shows the correlation between latent brain and behavior scores. Panel G illustrates OFC clusters that contributed significantly to LV2, with bootstrap ratios thresholded at ±1.96. Panel H shows Spearman correlations between oddball behavioral measures and LV2 brain scores. All red dashed lines mark the correlation threshold for *p* = .05. PLSC was conducted using post–pre difference scores for both OFC volume and oddball behavioral measures. Abbreviations: OFC = orbitofrontal cortex; RT = reaction time; PDR = pupil dilation response; P3 amplitude/latency at Fz = P300 amplitude/latency at frontal midline electrode; target/distractor = oddball task condition.

For LV1, the explained-variance profile showed that it accounted for the largest share of cross-block covariance (Figure 5B). The latent brain and behavioral scores were significantly correlated (*r* = .525, *p* < .001; Figure 5C). On the brain side, contributions to LV1 were localized within the two left OFC ROIs, with absolute bootstrap ratios exceeding 1.96 (Figure 5D). As shown in the figure, portions of both clusters contributed to the brain latent variable, indicating that structural increases in these OFC regions were most strongly associated with the LV1 pattern. On the behavioral side, two distractor-related measures showed significant associations with the brain latent score (Figure 5E). For P300 latency to distractor, increases in OFC cluster volume were associated with slower latencies. For pupil dilation to distractors, increases in OFC cluster volume were associated with reduced pupil responses. Thus, the associations indicate less rapid and robust response to task-irrelevant distractors. Although non-significant, reaction times to both distractor and target stimuli covaried in the opposite direction, suggesting that greater OFC cluster volume increases were associated with faster responses in general.

For LV2, the latent brain and behavioral scores were also significantly correlated (*r* = .541, *p* < .001; Figure 5F). A set of voxels within the left OFC clusters contributed significantly to this component, with absolute bootstrap ratios exceeding 1.96 (Figure 5G). On the behavioral side, target-related measures were most strongly associated with LV2 brain scores (Figure 5H). Specifically, increases in OFC volume were linked to smaller P300 amplitudes at Fz to both target and distractor stimuli. Because LV2 explained a smaller portion of cross-block covariance and was identified at a higher permutation *p*-value compared to LV1, this component should be interpreted with caution.

In summary, the voxelwise PLSC analysis revealed two significant latent variables linking OFC volume changes with oddball task performance. LV1 was primarily associated with distractor-related measures, including prolonged P300 latency, reduced pupil responses, and faster reaction times to both distractor and target stimuli. LV2 was characterized by reduced P300 amplitudes across both target- and distractor-related trials. These results indicate that OFC structural plasticity was related to multiple dimensions of task-evoked behavioral and physiological responses (Figure 5).

### 3.5. Brain–behavior associations between OFC volume changes and cognitive and neuropsychological task performance changes

In a separate voxelwise PLSC analysis, we examined whether OFC structural changes were linked to performance across a broader set of cognitive tasks. This analysis identified two significant latent variables (LV1, permutation *p* = .010; LV2, permutation *p* = .022), each capturing reliable associations between OFC volume changes and cognitive outcomes (Figure 6).

**Figure 6.**
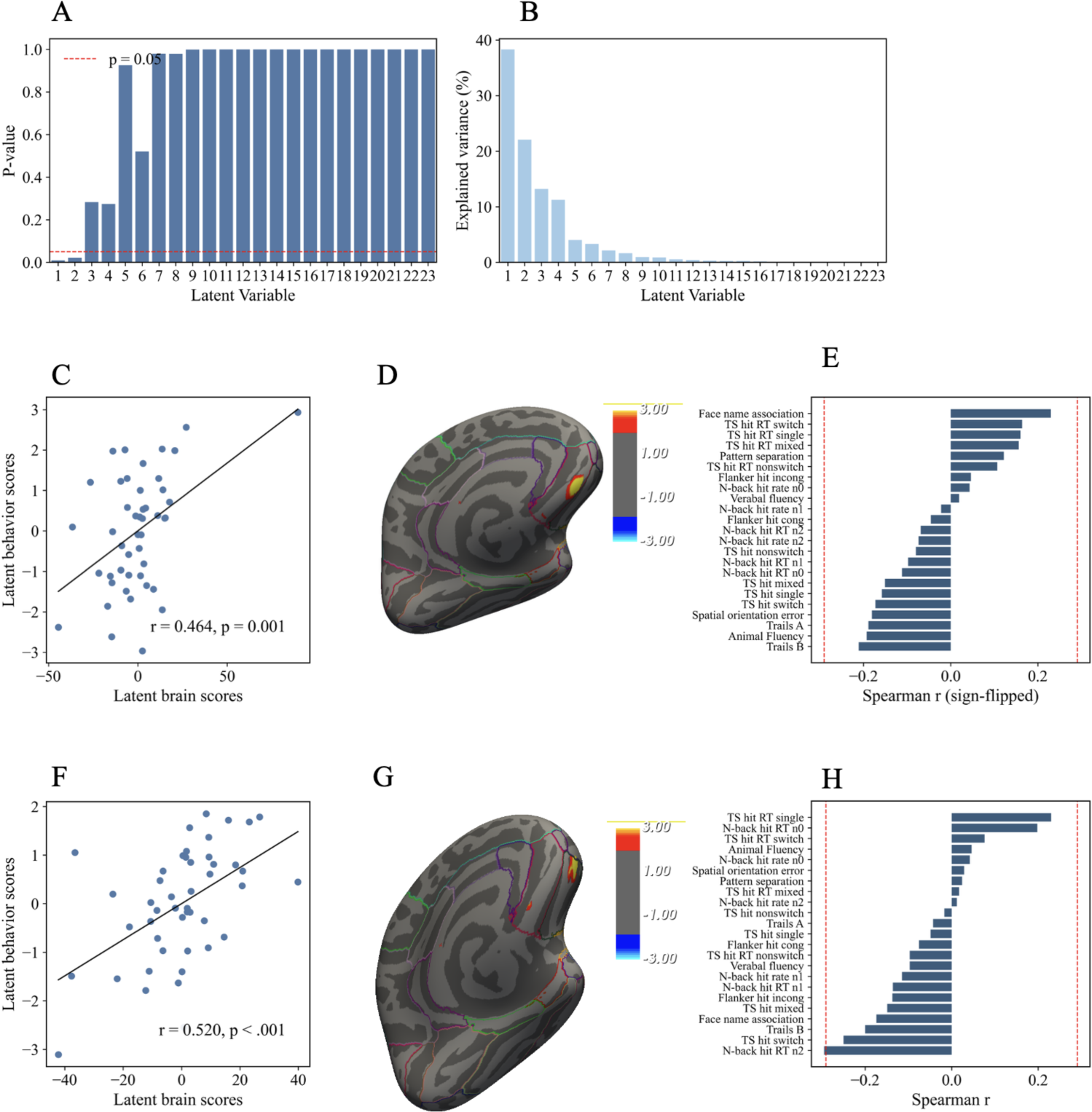
Vertexwise partial least squares correlation (PLSC) results: OFC volume change associated with cognitive task performance change. *Note*. Panels A–B show significance testing of latent variables. Panel A displays permutation p-values of each latent variable (LV). Panel B shows the explained variance (%) for each LV. Panels C–E present results for LV1. Panel C shows the correlation between latent brain and behavior scores. Panel D illustrates OFC clusters that contributed significantly to LV1, with bootstrap ratios thresholded at ±1.96. Panel E shows Spearman correlations between cognitive task measures and LV1 brain scores. Panels F–H present results for LV2. Panel F shows the correlation between latent brain and behavior scores. Panel G illustrates OFC clusters that contributed significantly to LV2, with bootstrap ratios thresholded at ±1.96. Panel H shows Spearman correlations between cognitive task measures and LV2 brain scores. All red dashed lines mark the correlation threshold for *p* = .05. PLSC was conducted using post–pre difference scores for both OFC volume and cognitive task measures. Abbreviations: OFC = orbitofrontal cortex; RT = reaction time; TS = task switching; Flanker = flanker task; N-back = N-back working memory task.

For LV1, the latent brain and behavioral scores were significantly correlated (*r* = .464, *p* = .001; Figure 6C). On the brain side, contributions to LV1 were localized within a medial OFC cluster, with absolute bootstrap ratios exceeding 1.96 (Figure 6D). As shown in the figure, this cluster contributed positively to the brain latent variable. On the behavioral side, most measures did not reach conventional significance, but several notable associations emerged (Figure 6E). Measures that covaried in the same direction as the medial OFC cluster included face–name associative memory, such that increases in OFC volume were linked to improved memory performance. In contrast, measures such as animal fluency and Trails A and B covaried in the opposite direction, suggesting that increases in OFC volume were associated with poorer performance on these tasks.

For LV2, the latent brain and behavioral scores were also significantly correlated (*r* = .520, *p* < .001; Figure 6F). A distinct cluster within the lateral OFC contributed positively to this component, with absolute bootstrap ratios exceeding 1.96 (Figure 6G). On the behavioral side, N-back reaction time showed an opposite relationship relative to the lateral OFC cluster, indicating that increases in lateral OFC volume were linked to faster responses. In contrast, task switching reaction times covaried in the same direction, such that increases in lateral OFC volume were associated with slower responses (Figure 6H).

In summary, the voxelwise PLSC analysis revealed two significant latent variables linking OFC volume changes with cognitive task performance. LV1 was driven by positive contributions from medial OFC voxels and showed mixed behavioral associations, including improved face–name memory but reduced fluency and trail-making performance. LV2 was characterized by positive contributions from lateral OFC voxels and showed opposing associations across tasks, with faster N-back responses but slower task switching responses. Together, these findings highlight anatomically distinct contributions of medial versus lateral OFC plasticity to multiple aspects of cognitive performance (Figure 6).

### 3.6. Brain–behavior associations between OFC volume changes and Lumosity game performance changes

For the Lumosity training outcomes, the voxelwise PLSC identified one significant latent variable (LV1, permutation *p* = .001), which accounted for the largest proportion of cross-block covariance (Figure 7A–B). The latent brain and behavioral scores were moderately and significantly correlated (*r* = .472, *p* < .001; Figure 7C).

**Figure 7.**
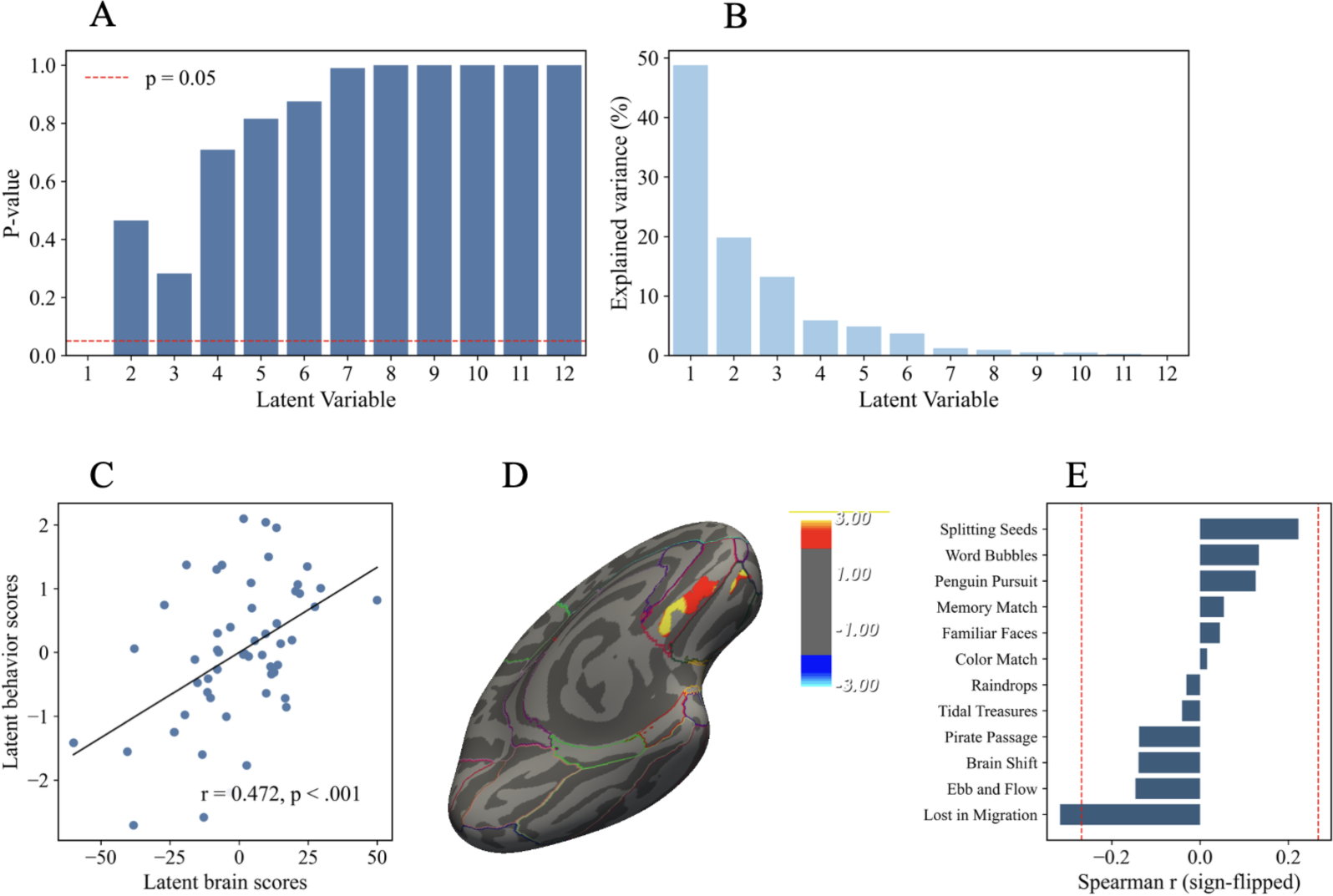
**Vertexwise partial least squares correlation (PLSC) results: OFC volume change associated with Lumosity game performance change** *Note*. Panels A–B show significance testing of latent variables. Panel A displays permutation *p*-values of each latent variable (LV), with the red dashed line indicating *p* = .05. Panel B shows the explained variance (%) for each LV. Panel C presents the correlation between latent brain and behavioral scores for LV1, with a least-squares fit; the annotation indicates *r* and *p*. Panel D illustrates OFC clusters that contributed significantly to LV1, with bootstrap ratios thresholded at ±1.96. Panel E shows Spearman correlations between individual Lumosity game measures and LV1 brain scores, with red dashed lines indicating the correlation threshold for *p* = .05. PLSC was conducted using post–pre difference scores for both OFC volume and Lumosity game measures. Abbreviations: OFC = orbitofrontal cortex. Lumosity games include: Lost in Migration, Ebb and Flow, Brain Shift, Pirate Passage, Tidal Treasures, Raindrops, Color Match, Familiar Faces, Memory Match, Penguin Pursuit, Word Bubbles, and Splitting Seeds.

On the brain side, contributions to LV1 were localized within the left OFC clusters, with absolute bootstrap ratios exceeding 1.96 (Figure 7D). These contributions were positive, indicating that structural increases in OFC volume were most strongly associated with the LV1 pattern.

On the behavioral side, several games, including Splitting Seeds, Word Bubbles, and Penguin Pursuit, covaried in the same direction as the OFC clusters (Figure 7E). By contrast, Lost in Migration covaried in the opposite direction, such that greater OFC volume increases were associated with reduced efficiency in this task.

Taken together, these results suggest that HRV-related structural changes in the left OFC were differentially related to gamified cognitive performance, with consistent same-direction associations for attention- and memory-oriented games, but opposite-direction associations for tasks requiring conflict monitoring under speeded conditions.

## 4. Discussion

The present study investigated how HRV biofeedback training relates to structural change in the OFC and to behavioral outcomes across multiple domains. In ROI-based analyses, HRV power during training, particularly within the upper LF band, was positively correlated with pre–post differences in the left lateral and medial OFC volumes. Consistent with these findings, vertex-wise analyses also revealed two clusters within these ROIs that showed significant positive correlations. Building on this, we carried out vertexwise PLSC restricted to these two left OFC clusters, which revealed linkages between changes in volume and changes in oddball performance, broader cognitive measures, and Lumosity training performance.

These patterns are consistent with our previous cross-sectional findings identifying the left OFC as the cortical region most reliably associated with HRV (Koenig et al., 2021; Yoo et al., 2018), as well as our findings that, in a randomized controlled trial, five weeks of HRV biofeedback training led to increased volume in both the lateral and medial OFC compared with an active control (Yoo et al., 2022). The present vertexwise findings reinforce and extend these findings by showing that training-induced structural changes in these same left OFC subdivisions are not only observable at the group level but also multivariately tied to performance changes.

Beyond this general pattern, the findings highlight the frequency specificity of the observed effects. The associations between HRV and OFC volume change were strongest near the upper low-frequency range, consistent with prior work highlighting resonance-frequency breathing around 0.1 Hz as optimal for engaging autonomic–prefrontal pathways (Lehrer & Gevirtz, 2014; Mather & Thayer, 2018). However, although the frequency-resolved analyses indicated that the peak effects occurred around 0.09 Hz, correlations extended into faster breathing-related frequencies, suggesting that the relevant physiological window may be broader than traditionally defined (Figure 4).

The vertex-wise PLSC within the OFC ROIs and oddball task measures identified two significant latent variables (LV1 and LV2), each reflecting different facets of the brain–behavior relationship. LV1 linked OFC volume increases with stronger attentional control over distractors, as evidenced by reduced pupil responses, faster reaction times, and prolonged P300 latencies—indicative of a shift toward a more accuracy-oriented, deliberate processing style. LV2, in contrast, linked OFC volume increases with reduced P300 amplitudes, pointing to a dampening of overall stimulus-driven reactivity, consistent with broader adjustments in regulatory control.

In the cognitive task domain, LV1 associated medial OFC volume increases with better associative memory but slower performance on speeded executive tasks, reflecting a speed–accuracy trade-off within executive control processes. LV2 highlighted contributions of the lateral OFC, where volume increases were associated with improved working memory efficiency (faster N-back responses) but impaired conflict monitoring (slower task switching responses), again indicating domain-specific trade-offs in executive control.

The Lumosity training results converged with these findings. OFC volume increases were linked to improved performance across several tasks that emphasize selective attention and working memory, but poorer performance on Lost in Migration, a task involving conflict monitoring, paralleling the PLSC results in the flanker task.

Together, these convergent findings suggest that OFC plasticity may facilitate attentional control and working memory while at times introducing trade-offs in conflict monitoring under speeded conditions. Recent work has emphasized functional heterogeneity within orbitofrontal subregions, demonstrating temporally distinct contributions of medial and lateral OFC circuits to autonomic and decision-related processes. Accordingly, the differential associations observed in the present study across medial and lateral OFC clusters may reflect distinct regulatory computations rather than uniform structural enhancement (Papageorgiou et al., 2026). Pupil dilation, as measured in the oddball task, has long been established as a sensitive index of attention allocation and arousal, reflecting activity of the LC–norepinephrine system (Murphy et al., 2011; Wainstein et al., 2017). Reduced pupil responses to distractors therefore may indicate diminished autonomic reactivity and more efficient suppression of irrelevant but salient stimuli. The OFC’s role in exerting top–down inhibitory control over distracting stimuli has been demonstrated in lesion and neurophysiological studies (Asko et al., 2024; Roberts & Wallis, 2000), and delayed P300 latency is often interpreted as indicative of more intentional, effortful stimulus evaluation rather than automatic orienting (Gazzaley et al., 2008; Huster et al., 2020). This alignment between our findings and the literature supports the interpretation that HRV-driven structural change in left OFC facilitates more efficient suppression and processing of distractors rather than accelerated processing per se.

In younger adults, higher resting HRV has been linked to better executive functioning, including inhibition, shifting, and working memory updating (Hansen et al., 2009; Williams et al., 2019). Evidence in older adults shows a similar relationship, though the relationship appears more heterogeneous, with age-related reductions in autonomic flexibility and greater inter-individual variability in health status likely contributing to variability in HRV–cognition associations (Forte et al., 2019; Kimhy et al., 2013). A systematic review concluded that reduced HRV, reflecting diminished parasympathetic activity, is broadly associated with poorer cognitive performance, with the strongest evidence in executive domains (Forte et al., 2019).

Neuroimaging evidence further indicates that task-induced reductions in HRV are coupled with changes in regional cerebral blood flow across medial prefrontal, anterior cingulate, and insular cortices, as well as subcortical structures, suggesting diminished prefrontal–autonomic regulation under greater cognitive demands in aging (Gianaros et al., 2004). Extending these findings, a randomized controlled trial showed that baseline executive functions explained significant variance in HRV among adults over 70 and that exergame-based cognitive–motor training selectively improved HRV indices (Eggenberger et al., 2020). In the present study, specific latent variables also indicated that increases in OFC volume were occasionally linked with slower responses in executive tasks. This pattern may reflect a speed–accuracy trade-off, particularly salient in older adults where limited cognitive resources necessitate prioritizing accuracy over speed. Evidence from younger adults also indicates that individuals with higher resting HRV perform more accurately but with slower reaction times on executive function tasks (Hansen et al., 2009). However, relationships between brain structure and behavior are not necessarily linear. Structural neuroimaging studies have emphasized that greater regional volume does not uniformly translate into better performance, as volumetric measures derived from voxel-based morphometry reflect a composite of multiple biological processes and may relate to behavior in developmentally and context-dependent ways (Kanai & Rees, 2011). From this perspective, the present findings may reflect a non-linear relationship in which OFC plasticity supports cognitive control up to an optimal range, beyond which additional structural engagement may introduce costs in processing speed or flexibility, particularly under speeded or conflict-laden task demands. Such effects may be especially pronounced in older adults, where compensatory recruitment of prefrontal control systems is common. These converging findings raise the possibility that the OFC structural changes induced by HRV biofeedback may promote a processing style that tends to emphasize accuracy over speed, particularly under cognitively demanding conditions. Such adaptations could encourage a strategically different mode of responding, which, depending on task demands and individual resource constraints, may manifest as slower reaction times in complex or speeded tasks.

Overall, these findings demonstrate the potential for HRV biofeedback to induce structural plasticity in prefrontal regions that support executive and attentional control, offering a mechanistic account of how autonomic regulation training may translate into cognitive benefits. Converging evidence further indicates that vagally mediated HRV covaries with distributed functional connectivity patterns across cortical networks in older adults, supporting the view that autonomic regulation is embedded within large-scale neural systems rather than isolated regions (Peralta-Malváez et al., 2023). From this perspective, the present OFC-specific structural changes may reflect a localized manifestation of broader autonomic–neural coupling processes. At the same time, the observed trade-offs indicate that such plasticity may not uniformly enhance performance across all domains, but instead may shift processing strategies toward accuracy-oriented control at the expense of speed or conflict monitoring.

Although the present sample focused on adults aged 50-70, an important open question is whether similar structural–behavioral adaptations would be observed in younger populations. Prior work suggests that higher resting HRV is linked to executive advantages across the lifespan, but the precise manifestations of HRV-related OFC plasticity may differ depending on age-related constraints on cognitive resources. Addressing this question will be critical for clarifying the generalizability of HRV biofeedback effects across developmental stages. By integrating neuroimaging, autonomic physiology, and behavioral measures, such work will be critical for advancing a mechanistic understanding of HRV biofeedback and its potential as a scalable intervention to support cognitive and emotional health across the lifespan.

## CRediT Authorship Contribution Statement

The authors made the following contributions. Hyun Joo Yoo: Conceptualization, Data curation, Formal analysis, Investigation, Methodology, Software, Visualization, Writing – original draft; Andy Jeesu Kim: Conceptualization, Data curation, Formal analysis, Investigation, Validation, Writing – review & editing; Martin J. Dahl: Conceptualization, Data curation, Methodology, Writing – review & editing; Kalekirstos Alemu: Investigation, Validation, Writing – review & editing; Kaoru Nashiro: Conceptualization, Data curation, Investigation, Project administration, Writing – review & editing; Christine Cho: Conceptualization, Data curation, Investigation, Resources, Project administration, Writing – review & editing; Noah Mercer: Data curation, Software, Investigation; Paul Choi: Data curation, Investigation, Project administration; Hye Rynn J. Lee: Data curation, Investigation, Project administration; Jungwon Min: Conceptualization, Data curation, Investigation; Julian Thayer: Conceptualization, Writing – review & editing; Mara Mather: Conceptualization, Methodology, Funding acquisition, Resources, Project administration, Supervision, Writing – review & editing.

## Data Availability Statement

The raw neuroimaging data and pulse data used for the analyses in this study will be made publicly available through OpenNeuro upon publication of this article. Additional behavioral data, processed and derived neuroimaging data, and physiological datasets used in the statistical analyses will also be provided via OpenNeuro as part of the curated dataset. All analysis code will be made publicly available upon publication through a GitHub repository. The permanent links to these repositories will be included in the final published version of the manuscript.

## References

1. Åhs, F., Sollers, J. J., III, Furmark, T., Fredrikson, M., & Thayer, J. F. (2009). High-frequency heart rate variability and cortico-striatal activity in men and women with social phobia. NeuroImage, 47(3), 815–820. 10.1016/j.neuroimage.2009.05.091

2. Angrilli, A., Palomba, D., Cantagallo, A., Maietti, A., & Stegagno, L. (1999). Emotional impairment after right orbitofrontal lesion in a patient without cognitive deficits. Neuroreport, 10(8), 1741–1746. 10.1097/00001756-199906030-00021

3. Asko, O., Blenkmann, A. O., Leske, S. L., Foldal, M. D., LLorens, A., Funderud, I., Meling, T. R., Knight, R. T., Endestad, T., & Solbakk, A.-K. (2024). Altered hierarchical auditory predictive processing after lesions to the orbitofrontal cortex. eLife, 13, e86386. 10.7554/eLife.86386

4. Aston-Jones, G., & Cohen, J. D. (2005). An integrative theory of locus coeruleus-norepinephrine function: adaptive gain and optimal performance. Annual Review of Neuroscience, 28(1), 403–450. 10.1146/annurev.neuro.28.061604.135709

5. Aston-Jones, G., Zhu, Y., & Card, J. P. (2004). Numerous GABAergic afferents to locus ceruleus in the pericerulear dendritic zone: possible interneuronal pool. The Journal of Neuroscience: The Official Journal of the Society for Neuroscience, 24(9), 2313–2321. 10.1523/JNEUROSCI.5339-03.2004

6. Beauchaine, T. P., & Thayer, J. F. (2015). Heart rate variability as a transdiagnostic biomarker of psychopathology. International Journal of Psychophysiology: Official Journal of the International Organization of Psychophysiology, 98(2 Pt 2), 338–350. 10.1016/j.ijpsycho.2015.08.004

7. Benjamini, Y., & Hochberg, Y. (1995). Controlling the false discovery rate: A practical and powerful approach to multiple testing. Journal of the Royal Statistical Society. Series B, Statistical Methodology, 57(1), 289–300. 10.1111/j.2517-6161.1995.tb02031.x

8. Brainard, D. H. (1997). The Psychophysics Toolbox. Spatial Vision, 10(4), 433–436. 10.1163/156856897x00357

9. Canning, S. J. D., Leach, L., Stuss, D., Ngo, L., & Black, S. E. (2004). Diagnostic utility of abbreviated fluency measures in Alzheimer disease and vascular dementia. Neurology, 62(4), 556–562. 10.1212/wnl.62.4.556

10. Chalmers, J. A., Quintana, D. S., Abbott, M. J.-A., & Kemp, A. H. (2014). Anxiety disorders are associated with reduced heart rate variability: A meta-analysis. Frontiers in Psychiatry, 5, 80. 10.3389/fpsyt.2014.00080

11. Chang, C., Metzger, C. D., Glover, G. H., Duyn, J. H., Heinze, H.-J., & Walter, M. (2013). Association between heart rate variability and fluctuations in resting-state functional connectivity. NeuroImage, 68, 93–104. 10.1016/j.neuroimage.2012.11.038

12. Critchley, H., Mathias, C., Josephs, O., O’Doherty, J., Zanini, S., Dewar, B., Cipolotti, L., Shallice, T., & Dolan, R. (2003). Human cingulate cortex and autonomic control: converging neuroimaging and clinical evidence. Brain : A Journal of Neurology, 126 *Pt* *10*, 2139–2152. 10.1093/BRAIN/AWG216

13. Dahl, M. J., Li, T., Mather, M., & Werkle-Bergner, M. (2025). Locus coeruleus-related insula activation supports implicit learning. In bioRxivorg (p. 2025.05.10.653174). 10.1101/2025.05.10.653174

14. Dale, A. M., Fischl, B., & Sereno, M. I. (1999). Cortical surface-based analysis. I. Segmentation and surface reconstruction. NeuroImage, 9(2), 179–194. 10.1006/nimg.1998.0395

15. Debnath, R., Buzzell, G. A., Morales, S., Bowers, M. E., Leach, S. C., & Fox, N. A. (2020). The Maryland analysis of developmental EEG (MADE) pipeline: The Maryland analysis of developmental EEG (MADE) pipeline. Psychophysiology, 57(6), e13580. 10.1111/psyp.13580

16. Delorme, A., & Makeig, S. (2004). EEGLAB: an open source toolbox for analysis of single-trial EEG dynamics including independent component analysis. Journal of Neuroscience Methods, 134(1), 9–21. 10.1016/j.jneumeth.2003.10.009

17. Desikan, R. S., Ségonne, F., Fischl, B., Quinn, B. T., Dickerson, B. C., Blacker, D., Buckner, R. L., Dale, A. M., Maguire, R. P., Hyman, B. T., Albert, M. S., & Killiany, R. J. (2006). An automated labeling system for subdividing the human cerebral cortex on MRI scans into gyral based regions of interest. NeuroImage, 31(3), 968–980. 10.1016/j.neuroimage.2006.01.021

18. Draheim, C., Tsukahara, J. S., Martin, J. D., Mashburn, C. A., & Engle, R. W. (2021). A toolbox approach to improving the measurement of attention control. Journal of Experimental Psychology. General, 150(2), 242–275. 10.1037/xge0000783

19. Eggenberger, P., Annaheim, S., Kündig, K. A., Rossi, R. M., Münzer, T., & de Bruin, E. D. (2020). Heart rate variability mainly relates to cognitive executive functions and improves through exergame training in older adults: A secondary analysis of a 6-month randomized controlled trial. Frontiers in Aging Neuroscience, 12, 197. 10.3389/fnagi.2020.00197

20. Eslinger, P. J., & Damasio, A. R. (1985). Severe disturbance of higher cognition after bilateral frontal lobe ablation: patient EVR. Neurology, 35(12), 1731–1741. 10.1212/wnl.35.12.1731

21. Fischl, B., Salat, D. H., Busa, E., Albert, M., Dieterich, M., Haselgrove, C., van der Kouwe, A., Killiany, R., Kennedy, D., Klaveness, S., Montillo, A., Makris, N., Rosen, B., & Dale, A. M. (2002). Whole brain segmentation. Neuron, 33(3), 341–355. 10.1016/s0896-6273(02)00569-x

22. Forte, G., Favieri, F., & Casagrande, M. (2019). Heart rate variability and cognitive function: A systematic review. Frontiers in Neuroscience, 13, 710. 10.3389/fnins.2019.00710

23. Friedman, A., Kohler, B., Gunalp, P., Boone, A. P., & Hegarty, M. (2020). A computerized spatial orientation test. Behavior Research Methods, 52(2), 799–812. 10.3758/s13428-019-01277-3

24. Fujii, T., Suzuki, M., Suzuki, K., Ohtake, H., Tsukiura, T., & Miura, R. (2005). Normal memory and no confabulation after extensive damage to the orbitofrontal cortex. *Journal of Neurology*, Neurosurgery, and Psychiatry, 76(9), 1309–1310. 10.1136/jnnp.2004.061846

25. Gajewski, P. D., & Falkenstein, M. (2012). Training-induced improvement of response selection and error detection in aging assessed by task switching: effects of cognitive, physical, and relaxation training. Frontiers in Human Neuroscience, 6, 130. 10.3389/fnhum.2012.00130

26. García Martínez, C. A., Otero Quintana, A., Vila, X. A., Lado Touriño, M. J., Rodríguez-Liñares, L., Rodríguez Presedo, J. M., & Méndez Penín, A. J. (2024). Heart rate variability analysis with the R package RHRV. Springer International Publishing. 10.1007/978-3-031-65753-5

27. Gazzaley, A., Clapp, W., Kelley, J., McEvoy, K., Knight, R. T., & D’Esposito, M. (2008). Age-related top-down suppression deficit in the early stages of cortical visual memory processing. Proceedings of the National Academy of Sciences of the United States of America, 105(35), 13122–13126. 10.1073/pnas.0806074105

28. Gianaros, P. J., Van Der Veen, F. M., & Jennings, J. R. (2004). Regional cerebral blood flow correlates with heart period and high-frequency heart period variability during working-memory tasks: Implications for the cortical and subcortical regulation of cardiac autonomic activity. Psychophysiology, 41(4), 521–530. 10.1111/1469-8986.2004.00179.x

29. Glasser, M. F., Sotiropoulos, S. N., Wilson, J. A., Coalson, T. S., Fischl, B., Andersson, J. L., Xu, J., Jbabdi, S., Webster, M., Polimeni, J. R., Van Essen, D. C., Jenkinson, M., & WU-Minn HCP Consortium. (2013). The minimal preprocessing pipelines for the Human Connectome Project. NeuroImage, 80, 105–124. 10.1016/j.neuroimage.2013.04.127

30. Glasser, M. F., & Van Essen, D. C. (2011). Mapping human cortical areas in vivo based on myelin content as revealed by T1- and T2-weighted MRI. The Journal of Neuroscience: The Official Journal of the Society for Neuroscience, 31(32), 11597–11616. 10.1523/JNEUROSCI.2180-11.2011

31. Goldstein, D. S., Bentho, O., Park, M.-Y., & Sharabi, Y. (2011). Low-frequency power of heart rate variability is not a measure of cardiac sympathetic tone but may be a measure of modulation of cardiac autonomic outflows by baroreflexes. Experimental Physiology, 96(12), 1255–1261. 10.1113/expphysiol.2010.056259

32. Gollan, T. H., Weissberger, G. H., Runnqvist, E., Montoya, R. I., & Cera, C. M. (2012). Self-ratings of spoken language dominance: A multi-lingual naming test (MINT) and preliminary norms for young and aging Spanish-English bilinguals. *Bilingualism (Cambridge*, England), 15(3), 594–615. 10.1017/S1366728911000332

33. Greve, D. N., & Fischl, B. (2009). Accurate and robust brain image alignment using boundary-based registration. NeuroImage, 48(1), 63–72. 10.1016/j.neuroimage.2009.06.060

34. Hansen, A. L., Johnsen, B. H., & Thayer, J. F. (2009). Relationship between heart rate variability and cognitive function during threat of shock. *Anxiety*, Stress, and Coping, 22(1), 77–89. 10.1080/10615800802272251

35. HeartMath Institute. (2020). emwave Pro [Computer software]. HeartMath Institute.

36. Herrera, A. Y., Velasco, R., Faude, S., White, J. D., Opitz, P. C., Huang, R., Tu, K., & Mather, M. (2020). Brain activity during a post-stress working memory task differs between the hormone-present and hormone-absent phase of hormonal contraception. Neurobiology of Stress, 13(100248), 100248. 10.1016/j.ynstr.2020.100248

37. Huster, R. J., Messel, M. S., Thunberg, C., & Raud, L. (2020). The P300 as marker of inhibitory control - Fact or fiction? Cortex; a Journal Devoted to the Study of the Nervous System and Behavior, 132, 334–348. 10.1016/j.cortex.2020.05.021

38. Jackson, A. S., Blair, S. N., Mahar, M. T., Wier, L. T., Ross, R. M., & Stuteville, J. E. (1990). Prediction of functional aerobic capacity without exercise testing. Medicine and Science in Sports and Exercise, 22(6), 863–870. 10.1249/00005768-199012000-00021

39. Jarczok, M. N., Jarczok, M., Mauss, D., Koenig, J., Li, J., Herr, R. M., & Thayer, J. F. (2013). Autonomic nervous system activity and workplace stressors--a systematic review. Neuroscience and Biobehavioral Reviews, 37(8), 1810–1823. 10.1016/j.neubiorev.2013.07.004

40. Jarczok, M. N., Weimer, K., Braun, C., Williams, D. P., Thayer, J. F., Gündel, H. O., & Balint, E. M. (2022). Heart rate variability in the prediction of mortality: A systematic review and meta-analysis of healthy and patient populations. Neuroscience and Biobehavioral Reviews, 143(104907), 104907. 10.1016/j.neubiorev.2022.104907

41. Jodo, E., & Aston-Jones, G. (1997). Activation of locus coeruleus by prefrontal cortex is mediated by excitatory amino acid inputs. Brain Research, 768(1-2), 327–332. 10.1016/s0006-8993(97)00703-8

42. Jodo, E., Chiang, C., & Aston-Jones, G. (1998). Potent excitatory influence of prefrontal cortex activity on noradrenergic locus coeruleus neurons. Neuroscience, 83(1), 63–79. 10.1016/s0306-4522(97)00372-2

43. Kanai, R., & Rees, G. (2011). The structural basis of inter-individual differences in human behaviour and cognition. Nature Reviews. Neuroscience, 12(4), 231–242. 10.1038/nrn3000

44. Kaufmann, T., Vögele, C., Sütterlin, S., Lukito, S., & Kübler, A. (2012). Effects of resting heart rate variability on performance in the P300 brain-computer interface. International Journal of Psychophysiology: Official Journal of the International Organization of Psychophysiology, 83(3), 336–341. 10.1016/j.ijpsycho.2011.11.018

45. Kim, A. J., Morales, S., Senior, J., & Mather, M. (2026). Electroencephalography, pupillometry, and behavioral evidence for locus coeruleus-noradrenaline system related tonic hyperactivity in older adults. Neurobiology of Aging, 159, 15–32. 10.1016/j.neurobiolaging.2025.11.008

46. Kimhy, D., Crowley, O. V., McKinley, P. S., Burg, M. M., Lachman, M. E., Tun, P. A., Ryff, C. D., Seeman, T. E., & Sloan, R. P. (2013). The association of cardiac vagal control and executive functioning--findings from the MIDUS study. Journal of Psychiatric Research, 47(5), 628–635. 10.1016/j.jpsychires.2013.01.018

47. Koenig, J., Abler, B., Agartz, I., Åkerstedt, T., Andreassen, O. A., Anthony, M., Bär, K.-J., Bertsch, K., Brown, R. C., Brunner, R., Carnevali, L., Critchley, H. D., Cullen, K. R., de Geus, E. J. C., de la Cruz, F., Dziobek, I., Ferger, M. D., Fischer, H., Flor, H., … Quintana, D. S. (2021). Cortical thickness and resting-state cardiac function across the lifespan: A cross-sectional pooled mega-analysis. Psychophysiology, 58(7), e13688. 10.1111/psyp.13688

48. Kok, A. (2001). On the utility of P3 amplitude as a measure of processing capacity. Psychophysiology, 38(3), 557–577. 10.1017/S0048577201990559

49. Krishnan, A., Williams, L. J., McIntosh, A. R., & Abdi, H. (2011). Partial Least Squares (PLS) methods for neuroimaging: a tutorial and review. NeuroImage, 56(2), 455–475. 10.1016/j.neuroimage.2010.07.034

50. Kromenacker, B. W., Sanova, A. A., Marcus, F. I., Allen, J. J. B., & Lane, R. D. (2018). Vagal mediation of low-frequency heart rate variability during slow yogic breathing. Psychosomatic Medicine, 80(6), 581–587. 10.1097/PSY.0000000000000603

51. Kuo, C.-C., Hsieh, J.-C., Tsai, H.-C., Kuo, Y.-S., Yau, H.-J., Chen, C.-C., Chen, R.-F., Yang, H.-W., & Min, M.-Y. (2020). Inhibitory interneurons regulate phasic activity of noradrenergic neurons in the mouse locus coeruleus and functional implications. The Journal of Physiology, 598(18), 4003–4029. 10.1113/JP279557

52. Lane, R. D., McRae, K., Reiman, E. M., Chen, K., Ahern, G. L., & Thayer, J. F. (2009). Neural correlates of heart rate variability during emotion. NeuroImage, 44(1), 213–222. 10.1016/j.neuroimage.2008.07.056

53. Leach, S. C., Morales, S., Bowers, M. E., Buzzell, G. A., Debnath, R., Beall, D., & Fox, N. A. (2020). Adjusting ADJUST: Optimizing the ADJUST algorithm for pediatric data using geodesic nets. Psychophysiology, 57(8), e13566. 10.1111/psyp.13566

54. Lehrer, P. (2013). How does heart rate variability biofeedback work? Resonance, the baroreflex, and other mechanisms. Biofeedback, 41(1), 26–31. https://meridian.allenpress.com/biofeedback/article-abstract/41/1/26/113345

55. Lehrer, P., & Gevirtz, R. (2014). Heart rate variability biofeedback: how and why does it work? Frontiers in Psychology, 5, 756. 10.3389/fpsyg.2014.00756

56. Mather, M. (2020). The locus coeruleus-norepinephrine system role in cognition and how it changes with aging. In The Cognitive Neurosciences (pp. 91–104). The MIT Press.

57. Mather, M., Huang, R., Clewett, D., Nielsen, S. E., Velasco, R., Tu, K., Han, S., & Kennedy, B. L. (2020). Isometric exercise facilitates attention to salient events in women via the noradrenergic system. NeuroImage, 210(116560), 116560. 10.1016/j.neuroimage.2020.116560

58. Mather, M., & Thayer, J. (2018). How heart rate variability affects emotion regulation brain networks. Current Opinion in Behavioral Sciences, 19, 98–104. 10.1016/j.cobeha.2017.12.017

59. Matthews, S. C., Paulus, M. P., Simmons, A. N., Nelesen, R. A., & Dimsdale, J. E. (2004). Functional subdivisions within anterior cingulate cortex and their relationship to autonomic nervous system function. NeuroImage, 22(3), 1151–1156. 10.1016/j.neuroimage.2004.03.005

60. Matusik, P. S., Zhong, C., Matusik, P. T., Alomar, O., & Stein, P. K. (2023). Neuroimaging studies of the neural correlates of heart rate variability: A systematic review. Journal of Clinical Medicine, 12(3), 1016. 10.3390/jcm12031016

61. Menuet, C., Ben-Tal, A., Linossier, A., Allen, A. M., Machado, B. H., Moraes, D. J. A., Farmer, D. G. S., Paterson, D. J., Mendelowitz, D., Lakatta, E. G., Taylor, E. W., Ackland, G. L., Zucker, I. H., Fisher, J. P., Schwaber, J. S., Shanks, J., Paton, J. F. R., Buron, J., Spyer, K. M., … Gourine, A. V. (2025). Redefining respiratory sinus arrhythmia as respiratory heart rate variability: an international Expert Recommendation for terminological clarity. Nature Reviews. Cardiology. 10.1038/s41569-025-01160-z

62. Morales, S., Bowers, M. E., Leach, S. C., Buzzell, G. A., Fifer, W., Elliott, A. J., & Fox, N. A. (2022). Time-frequency dynamics of error monitoring in childhood: An EEG study. Developmental Psychobiology, 64(3), e22215. 10.1002/dev.22215

63. Murphy, P. R., O’Connell, R. G., O’Sullivan, M., Robertson, I. H., & Balsters, J. H. (2014). Pupil diameter covaries with BOLD activity in human locus coeruleus: Pupil Diameter and Locus Coeruleus Activity. Human Brain Mapping, 35(8), 4140–4154. 10.1002/hbm.22466

64. Murphy, P. R., Robertson, I. H., Balsters, J. H., & O’connell, R. G. (2011). Pupillometry and P3 index the locus coeruleus-noradrenergic arousal function in humans: Indirect markers of locus coeruleus activity. Psychophysiology, 48(11), 1532–1543. 10.1111/j.1469-8986.2011.01226.x

65. Namiki, C., Yamada, M., Yoshida, H., Hanakawa, T., Fukuyama, H., & Murai, T. (2008). Small orbitofrontal traumatic lesions detected by high resolution MRI in a patient with major behavioural changes. Neurocase, 14(6), 474–479. 10.1080/13554790802459494

66. Nashiro, K., Min, J., Yoo, H. J., Cho, C., Dahl, M. J., Choi, P., Lee, H. R. J., Choupan, J., Mercer, N., Nasseri, P., Kim, A. J., Alemu, K., Rose, N. F., Herrera, A. Y., Custer, R., Werkle-Bergner, M., Thayer, J. F., Sordo, L., Head, E., & Mather, M. (2026). Testing effects of paced breathing on plasma Aβ and brain perivascular spaces. International Journal of Psychophysiology: Official Journal of the International Organization of Psychophysiology, 221(113334), 113334. 10.1016/j.ijpsycho.2026.113334 (In press)

67. Nashiro, K., Yoo, H. J., Cho, C., Kim, A. J., Nasseri, P., Min, J., Dahl, M. J., Mercer, N., Choupan, J., Choi, P., Lee, H. R. J., Choi, D., Alemu, K., Herrera, A. Y., Ng, N. F., Thayer, J. F., & Mather, M. (2024). Heart rate and breathing effects on attention and memory (HeartBEAM): study protocol for a randomized controlled trial in older adults. Trials, 25(1), 190. 10.1186/s13063-024-07943-y

68. Nasreddine, Z. S., Phillips, N. A., Bédirian, V., Charbonneau, S., Whitehead, V., Collin, I., Cummings, J. L., & Chertkow, H. (2005). The Montreal Cognitive Assessment, MoCA: a brief screening tool for mild cognitive impairment: Moca: A brief screening tool for MCI. Journal of the American Geriatrics Society, 53(4), 695–699. 10.1111/j.1532-5415.2005.53221.x

69. Nieuwenhuis, S., Aston-Jones, G., & Cohen, J. (2005). Decision making, the P3, and the locus coeruleus-norepinephrine system. Psychological Bulletin, 131(4), 510–532. 10.1037/0033-2909.131.4.510

70. Nolan, H., Whelan, R., & Reilly, R. B. (2010). *FASTER*: Fully Automated Statistical Thresholding for EEG artifact Rejection. Journal of Neuroscience Methods, 192(1), 152–162. 10.1016/j.jneumeth.2010.07.015

71. Olfers, K. J. F., & Band, G. P. H. (2018). Game-based training of flexibility and attention improves task-switch performance: near and far transfer of cognitive training in an EEG study. Psychological Research, 82(1), 186–202. 10.1007/s00426-017-0933-z

72. Ottesen, J. T. (1997). Modelling of the baroreflex-feedback mechanism with time-delay. Journal of Mathematical Biology, 36(1), 41–63. 10.1007/s002850050089

73. Papageorgiou, G. K., Amemori, K.-I., Gibson, D. J., Schwerdt, H. N., Naim, M., Wang, M. C., Yoshida, T., Sharma, J., Upadhyay, U., Yang, G. R., & Graybiel, A. M. (2026). Functional distinctions between orbitofrontal cortex and anterior cingulate cortex subregions in decision-making and autonomic regulation. Nature Communications, 1–21. 10.1038/s41467-026-69447-4

74. Park, G., & Thayer, J. F. (2014). From the heart to the mind: cardiac vagal tone modulates top-down and bottom-up visual perception and attention to emotional stimuli. Frontiers in Psychology, 5, 278. 10.3389/fpsyg.2014.00278

75. Peralta-Malváez, L., Turnbull, A., Anthony, M., Adeli, E., & Lin, F. V. (2023). CCA identifies a neurophysiological marker of adaptation capacity that is reliably linked to internal locus of control of cognition in amnestic MCI. GeroScience, 45(3), 1803–1815. 10.1007/s11357-023-00730-8

76. Pfurtscheller, G., Schwerdtfeger, A., Seither-Preisler, A., Brunner, C., Aigner, C. S., Calisto, J., Gens, J., & Andrade, A. (2018). Synchronization of intrinsic 0.1-Hz blood-oxygen-level-dependent oscillations in amygdala and prefrontal cortex in subjects with increased state anxiety. The European Journal of Neuroscience, 47(5), 417–426. 10.1111/ejn.13845

77. Polcher, A., Frommann, I., Koppara, A., Wolfsgruber, S., Jessen, F., & Wagner, M. (2017). Face-name associative recognition deficits in subjective cognitive decline and mild cognitive impairment. Journal of Alzheimer’s Disease: JAD, 56(3), 1185–1196. 10.3233/JAD-160637

78. Polich, J. (2007). Updating P300: an integrative theory of P3a and P3b. Clinical Neurophysiology: Official Journal of the International Federation of Clinical Neurophysiology, 118(10), 2128–2148. 10.1016/j.clinph.2007.04.019

79. Reuter, M., Schmansky, N. J., Rosas, H. D., & Fischl, B. (2012). Within-subject template estimation for unbiased longitudinal image analysis. NeuroImage, 61(4), 1402–1418. 10.1016/j.neuroimage.2012.02.084

80. Reyes del Paso, G. A., Langewitz, W., Mulder, L. J. M., van Roon, A., & Duschek, S. (2013). The utility of low frequency heart rate variability as an index of sympathetic cardiac tone: a review with emphasis on a reanalysis of previous studies: LF HRV and sympathetic cardiac tone. Psychophysiology, 50(5), 477–487. 10.1111/psyp.12027

81. Ritz, T. (2024). Putting back respiration into respiratory sinus arrhythmia or high-frequency heart rate variability: Implications for interpretation, respiratory rhythmicity, and health. Biological Psychology, 185(108728), 108728. 10.1016/j.biopsycho.2023.108728

82. Roberts, A. C., & Wallis, J. (2000). Inhibitory control and affective processing in the prefrontal cortex: neuropsychological studies in the common marmoset. Cerebral Cortex (New York, N.Y.: 1991), 10(3), 252–262. 10.1093/CERCOR/10.3.252

83. Rodríguez-Liñares, L., Méndez, A. J., Lado, M. J., Olivieri, D. N., Vila, X. A., & Gómez-Conde, I. (2011). An open source tool for heart rate variability spectral analysis. Computer Methods and Programs in Biomedicine, 103(1), 39–50. 10.1016/j.cmpb.2010.05.012

84. Rossetti, H. C., Lacritz, L. H., Cullum, C. M., & Weiner, M. F. (2011). Normative data for the Montreal Cognitive Assessment (MoCA) in a population-based sample. Neurology, 77(13), 1272–1275. 10.1212/WNL.0b013e318230208a

85. Rudebeck, P. H., & Rich, E. L. (2018). Orbitofrontal cortex. Current Biology: CB, 28(18), R1083–R1088. 10.1016/j.cub.2018.07.018

86. Schneider, M., & Schwerdtfeger, A. (2020). Autonomic dysfunction in posttraumatic stress disorder indexed by heart rate variability: a meta-analysis. Psychological Medicine, 50, 1937–1948. 10.1017/S003329172000207X

87. Schwarzkopf, D. S., De Haas, B., & Rees, G. (2012). Better ways to improve standards in brain-behavior correlation analysis. Frontiers in Human Neuroscience, 6, 200. 10.3389/fnhum.2012.00200

88. Smith, A.-L., Owen, H., & Reynolds, K. J. (2015). Can short-term heart rate variability be used to monitor fentanyl-midazolam induced changes in ANS preceding respiratory depression? Journal of Clinical Monitoring and Computing, 29(3), 393–405. 10.1007/s10877-014-9617-z

89. Spreen, O., & Risser, A. H. (2003). *Assessment of aphasia*. https://books.google.com/books?hl=en&lr=&id=fBRnDAAAQBAJ&oi=fnd&pg=PR7&dq=Spreen+%26+Risser,+2003&ots=YAe64LIpXv&sig=q0EVBoMkUNgBsf4pHmc0-tfwZac

90. Steyvers, M., & Schafer, R. J. (2020). Inferring latent learning factors in large-scale cognitive training data. Nature Human Behaviour, 4(11), 1145–1155. 10.1038/s41562-020-00935-3

91. Stuss, D. T., Benson, D. F., Kaplan, E. F., Weir, W. S., Naeser, M. A., Lieberman, I., & Ferrill, D. (1983). The involvement of orbitofrontal cerebrum in cognitive tasks. Neuropsychologia, 21(3), 235–248. 10.1016/0028-3932(83)90040-4

92. Thayer, J. F., Hansen, A. L., Saus-Rose, E., & Johnsen, B. H. (2009). Heart rate variability, prefrontal neural function, and cognitive performance: the neurovisceral integration perspective on self-regulation, adaptation, and health. Annals of Behavioral Medicine: A Publication of the Society of Behavioral Medicine, 37(2), 141–153. 10.1007/s12160-009-9101-z

93. Thayer, J. F., & Lane, R. D. (2000). A model of neurovisceral integration in emotion regulation and dysregulation. Journal of Affective Disorders, 61(3), 201–216. 10.1016/s0165-0327(00)00338-4

94. Thayer, J. F., & Lane, R. D. (2009). Claude Bernard and the heart-brain connection: further elaboration of a model of neurovisceral integration. Neuroscience and Biobehavioral Reviews, 33(2), 81–88. 10.1016/j.neubiorev.2008.08.004

95. Thompson, P. M., Jahanshad, N., Ching, C. R. K., Salminen, L. E., Thomopoulos, S. I., Bright, J., Baune, B. T., Bertolín, S., Bralten, J., Bruin, W. B., Bülow, R., Chen, J., Chye, Y., Dannlowski, U., de Kovel, C. G. F., Donohoe, G., Eyler, L. T., Faraone, S. V., Favre, P., … ENIGMA Consortium. (2020). ENIGMA and global neuroscience: A decade of large-scale studies of the brain in health and disease across more than 40 countries. Translational Psychiatry, 10(1), 100. 10.1038/s41398-020-0705-1

96. Tombaugh, T. N. (2004). Trail Making Test A and B: normative data stratified by age and education. Archives of Clinical Neuropsychology: The Official Journal of the National Academy of Neuropsychologists, 19(2), 203–214. 10.1016/S0887-6177(03)00039-8

97. Vaschillo, E. G., Vaschillo, B., & Lehrer, P. (2006). Characteristics of resonance in heart rate variability stimulated by biofeedback. Applied Psychophysiology and Biofeedback, 31(2), 129–142. 10.1007/s10484-006-9009-3

98. Vaschillo, E. G., Vaschillo, B., Pandina, R. J., & Bates, M. E. (2011). Resonances in the cardiovascular system caused by rhythmical muscle tension. Psychophysiology, 48(7), 927–936. 10.1111/j.1469-8986.2010.01156.x

99. Vazey, E. M., Moorman, D. E., & Aston-Jones, G. (2018). Phasic locus coeruleus activity regulates cortical encoding of salience information. Proceedings of the National Academy of Sciences of the United States of America, 115(40), E9439–E9448. 10.1073/pnas.1803716115

100. Viola, F. C., Debener, S., Thorne, J., & Schneider, T. R. (2010). Using ICA for the analysis of multi-channel EEG data. *Simultaneous EEG and fMRI: Recording, Analysis, and Application: Recording*, Analysis, and Application, 121–133.

101. Wainstein, G., Rojas-Líbano, D., Rojas-Líbano, D., Crossley, N., Crossley, N., Carrasco, X., Aboitiz, F., & Ossandón, T. (2017). Pupil size tracks attentional performance in attention-deficit/hyperactivity disorder. Scientific Reports, 7. 10.1038/s41598-017-08246-w

102. Wais, P. E., Arioli, M., Anguera-Singla, R., & Gazzaley, A. (2021). Virtual reality video game improves high-fidelity memory in older adults. Scientific Reports, 11(1), 2552. 10.1038/s41598-021-82109-3

103. Williams, P. G., Cribbet, M. R., Tinajero, R., Rau, H. K., Thayer, J. F., & Suchy, Y. (2019). The association between individual differences in executive functioning and resting high-frequency heart rate variability. Biological Psychology, 148(107772), 107772. 10.1016/j.biopsycho.2019.107772

104. Yackle, K., Schwarz, L. A., Kam, K., Sorokin, J. M., Huguenard, J. R., Feldman, J. L., Luo, L., & Krasnow, M. A. (2017). Breathing control center neurons that promote arousal in mice. *Science (New York*, N.Y.), 355(6332), 1411–1415. 10.1126/science.aai7984

105. Yoo, H. J., Nashiro, K., Min, J., Cho, C., Bachman, S. L., Nasseri, P., Porat, S., Dutt, S., Grigoryan, V., Choi, P., Thayer, J. F., Lehrer, P. M., Chang, C., & Mather, M. (2022). Heart rate variability (HRV) changes and cortical volume changes in a randomized trial of five weeks of daily HRV biofeedback in younger and older adults. International Journal of Psychophysiology: Official Journal of the International Organization of Psychophysiology, 181, 50–63. 10.1016/j.ijpsycho.2022.08.006

106. Yoo, H. J., Thayer, J. F., Greening, S., Lee, T.-H., Ponzio, A., Min, J., Sakaki, M., Nga, L., Mather, M., & Koenig, J. (2018). Brain structural concomitants of resting state heart rate variability in the young and old: evidence from two independent samples. Brain Structure & Function, 223(2), 727–737. 10.1007/s00429-017-1519-7

