## Supplementary Materials for "Daily Paced Breathing Sessions Induce Left Orbitofrontal Volume Changes Linked to Cognitive Outcomes"

**Supplementary Figure S1**

*Overview of the Study Design and Analytical Framework Linking Training HRV, Cortical Plasticity, and Cognitive Performance*

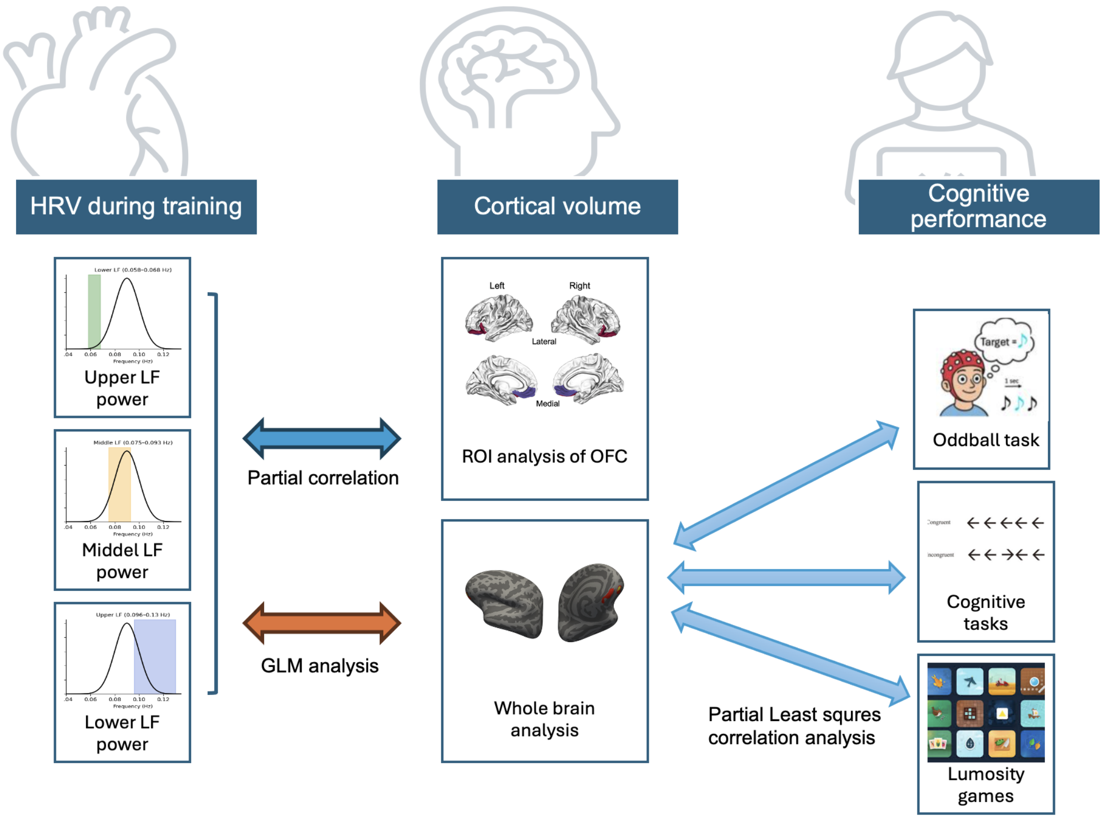


*Note.* The illustration of the auditory oddball task is adapted in part from (Kim et al., 2026). The Lumosity game image is reproduced from publicly available materials on the Lumosity website for illustrative purposes only.

**Supplementary Figure S2**

*Overview of the 12-week study schedule*

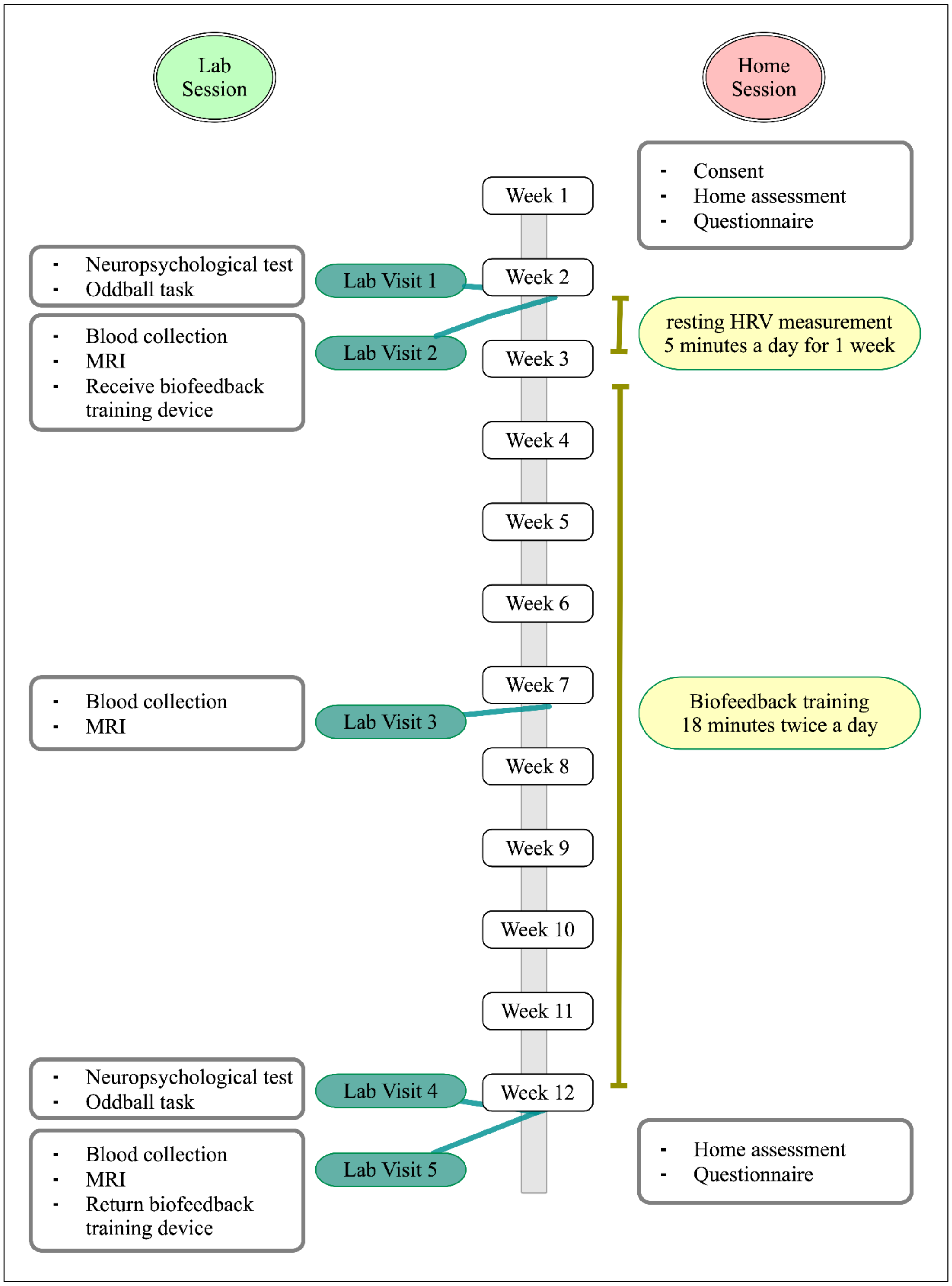

**Supplementary Figure S3**

*Visual breathing pacer interface used during training*

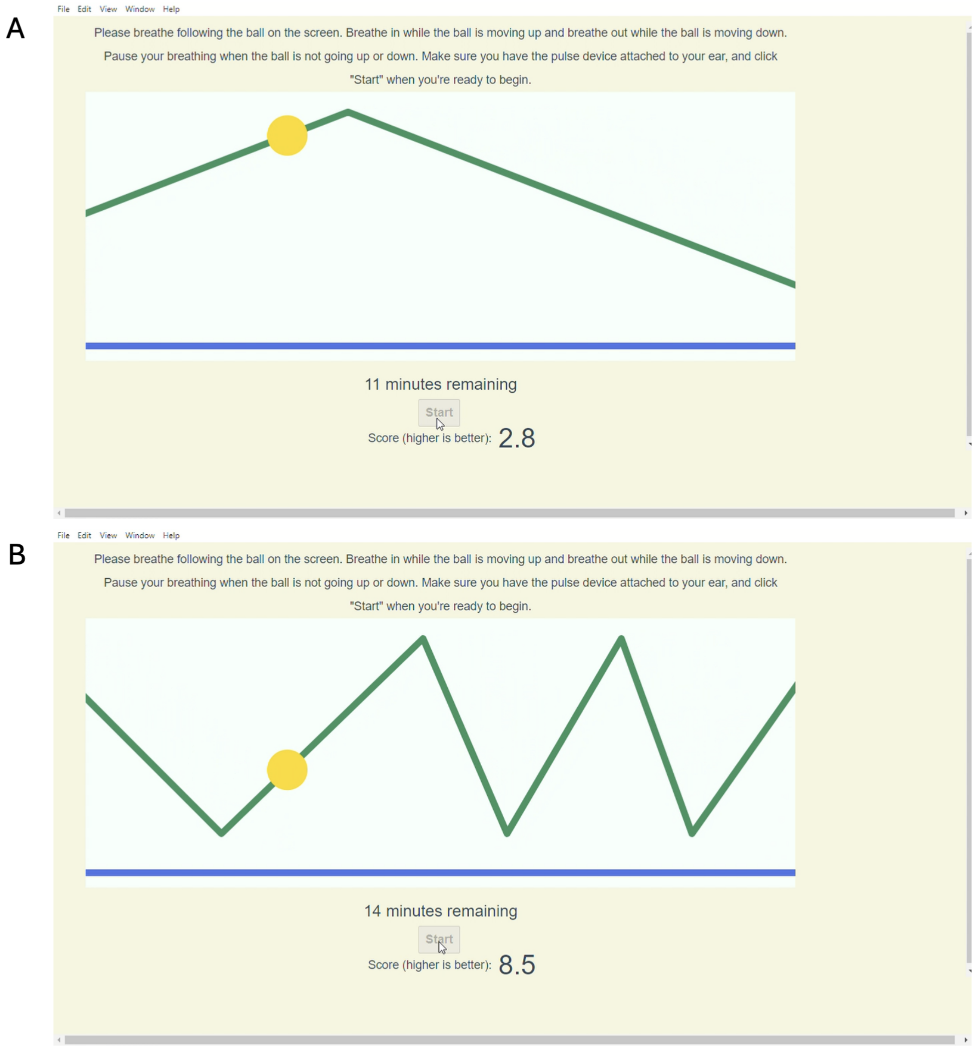


*Note.* Panels A and B show screenshots from the breathing training application used in the study. The application displayed a visual pacer consisting of a ball moving vertically along a line, which participants followed by inhaling as the ball moved upward and exhaling as it moved downward. Participants completed paced breathing segments according to their assigned condition (A: slow-paced breathing; B: random-paced breathing; see Methods).
During each segment, a condition-specific performance score was displayed below the pacer. In the slow-paced breathing condition, the score reflected the magnitude of heart rate oscillations, with higher values indicating greater oscillatory activity. In the random-paced breathing condition, the displayed score was inversely scaled such that higher values corresponded to reduced heart rate oscillations.

**Supplementary Table S1**

*Schedule of home-based cognitive tasks administered during week 1 and week 12.*

| Type | Test | Day 1 | Day 2 | Day 3 | Day 4 | Day 5 | Day 6 |
|---|---|---|---|---|---|---|---|
| Cognition | CVLT Part 1 | | | | | O | |
| | CVLT Part 2 (after 20 min interval) | | | | | O | |
| | Working memory (n-back) | | O | O | O | O | O |
| | Face-name associative memory | O | O | O | O | O | O |
| | Pattern separation task - Part 1 (encoding) | | O | | O | | O |
| | Pattern separation task - Part 2 (test) | | O | | O | | O |
| | Task switching | | | O | | | |
| | Flanker | | | O | | O | |
| | Spatial orientation | O | O | O | O | | O |
| | Verbal Fluency | O | O | O | O | | O |

*Note.* The same sequence of tasks was administered during Week 2. Empty cells indicate that the task was not administered on that day.

**Supplementary Table S2**

*Lumosity games*

| Set | Game name | Category | Description |
|---|---|---|---|
| 1 | Familiar faces | Memory | Participants play a waiter's role and earn higher tips if they can remember their customers' names and food orders |
| 1 | Tidal treasures | Memory | On each trial, participants are shown several unique ocean treasures and must choose one that they have not already selected in that round. Rounds can include up to 35 items, and some items are quite similar |
| 1 | Lost in migration | Attention | Participants indicate the direction of the central bird in the formation while ignoring the distractors around it |
| 1 | Splitting seeds | Attention | Participants evenly divide a pile of seeds without counting them |
| 1 | Pirate passage | Reasoning | Participants navigate their ship to reach the treasure island without colliding with other pirate ships |
| 1 | Ebb and flow | Flexibility | Participants view green or orange leaves moving across a pond and indicate the direction of where green leaves are pointing or where orange leaves are moving |
| 2 | Memory match | Memory | Participants must quickly determine whether a flashcard symbol matches the one presented two items ago |
| 2 | Word bubbles rising | Language | Participants generate as many words as possible that start with the same starting letters (e.g., res, medi) within the time limit |
| 2 | Raindrops | Math | Participants perform each math problem inside each raindrop before it reaches the bottom of the screen. Math problems include addition, subtraction, multiplication, and division |
| 2 | Penguin pursuit | Attention | Participants guide a penguin through a maze to reach a reward of fish at the end before the other penguin does. When the maze rotates, participants must rotate their mental map of the maze and recalibrate the directions to get to the goal |
| 2 | Brain shift | Flexibility | Participants are shown a letter-number pair (e.g., 5E) on the top or the bottom card. If the letter-number pair shows up on top, participants indicate whether the number is even; if it appears at the bottom, participants indicate whether the letter is a vowel or not |
| 2 | Color match 2 | Flexibility | Participants indicate the color of a written word while ignoring the meaning of the word |

**Supplementary Table S3**

*Basic participant characteristics across conditions*

| | All | Slow-paced breathing | Random-paced breathing | Condition difference (*t or* $\chi^2$) | Condition difference (*p*) |
|---|---|---|---|---|---|
| N | 55 | 29 | 26 | | |
| Age | 60.1 (5.99) | 60.6 (5.58) | 59.6 (6.48) | 0.50 | .553 |
| Gender | 29 F/26 M | 15 F/14 M | 14 F/12 M | 0.02 | .875 |
| Education | 16.6 (2.36) | 15.9 (1.87) | 17.4 (2.62) | -2.50 | .016 |
| MoCA | 26.4 (2.24) | 26.19 (2.25) | 26.52 (2.25) | -0.54 | .595 |
| Race | | | | 1.99 | .737 |
| African American | 6 | 2 | 4 | | |
| Asian | 11 | 7 | 4 | | |
| Bi-racial | 5 | 3 | 2 | | |
| Caucasian | 30 | 16 | 14 | | |
| Other | 0 | 0 | 0 | | |
| Prefer not to state | 3 | 1 | 2 | | |

$^*p < 0.05$; $^{**}p < 0.01$; $^{***}p < 0.001$, 2-tailed. Note: Condition differences were statistically tested using t-tests for age and education and chi-square tests for gender and race.
